# Polygenic risk for immuno-metabolic markers and specific depressive symptoms: A multi-sample network analysis study

**DOI:** 10.1101/2021.01.07.20248981

**Authors:** Nils Kappelmann, Darina Czamara, Nicolas Rost, Sylvain Moser, Vanessa Schmoll, Lucia Trastulla, Jan Stochl, Susanne Lucae, CHARGE inflammation working group, Elisabeth B. Binder, Golam M. Khandaker, Janine Arloth

## Abstract

**Background:** About every fourth patient with major depressive disorder (MDD) shows evidence of systemic inflammation. Previous studies have shown inflammation-depression associations of multiple serum inflammatory markers and multiple specific depressive symptoms. It remains unclear, however, if these associations extend to genetic/lifetime predisposition to higher inflammatory marker levels and what role metabolic factors such as Body Mass Index (BMI) play. It is also unclear whether inflammation-symptom associations reflect direct or indirect associations, which can be disentangled using network analysis.

**Methods:** This study examined associations of polygenic risk scores (PRSs) for immuno-metabolic markers (C-reactive protein [CRP], interleukin [IL]-6, IL-10, tumour necrosis factor [TNF]-α, BMI) with seven depressive symptoms in one general population sample, the UK Biobank study (n=110,010), and two patient samples, the Munich Antidepressant Response Signature (MARS, n=1,058) and Sequenced Treatment Alternatives to Relieve Depression (STAR*D, n=1,143) studies. Network analysis was applied jointly for these samples using fused graphical least absolute shrinkage and selection operator (FGL) estimation as primary analysis and, individually, using unregularized model search estimation. Stability of results was assessed using bootstrapping and three consistency criteria were defined to appraise robustness and replicability of results across estimation methods, network bootstrapping, and samples.

**Results:** Network analysis results displayed to-be-expected PRS-PRS and symptom-symptom associations (termed edges), respectively, that were mostly positive. Using FGL estimation, results further suggested 28, 29, and six PRS-symptom edges in MARS, STAR*D, and UK Biobank samples, respectively. Unregularized model search estimation suggested three PRS-symptom edges in the UK Biobank sample. Applying our consistency criteria to these associations indicated that only the association of higher CRP PRS with greater changes in appetite fulfilled all three criteria.

Four additional associations fulfilled at least two consistency criteria; specifically, higher CRP PRS was associated with greater fatigue and reduced anhedonia, higher TNF-α PRS was associated with greater fatigue, and higher BMI PRS with greater changes in appetite and anhedonia. Associations of the BMI PRS with anhedonia, however, showed an inconsistent valence across estimation methods.

**Conclusions:** Genetic predisposition to higher systemic inflammatory markers are primarily associated with somatic/neurovegetative symptoms of depression such as changes in appetite and fatigue, consistent with previous studies based on circulating levels of inflammatory markers. We extend these findings by providing evidence that associations are direct (using network analysis) and extend to genetic predisposition to immuno-metabolic markers (using PRSs). Our findings can inform selection of patients with inflammation-related symptoms into clinical trials of immune-modulating drugs for MDD.

## INTRODUCTION

Recent findings suggest that every fourth patient with Major Depressive Disorder (MDD) shows evidence of systemic, low-grade inflammation as indicated by elevated (>3mg/L) C-reactive protein (CRP) concentrations (Osimo et al., 2019). This association has been supported by cross-sectional case-control studies synthesised in multiple meta-analyses (Dowlati et al., 2010; Goldsmith et al., 2016; Haapakoski et al., 2015; Howren et al., 2009; Köhler et al., 2017) as well as longitudinal studies (Khandaker et al., 2014; Lamers et al., 2020; Mac Giollabhui et al., 2020). Clinically, patients with evidence of inflammation do not respond as well to standard monoaminergic and psychotherapeutic treatments (Liu et al., 2020; Lopresti, 2017). These patients may, however, benefit from alternative treatment with immune-modulating drugs (Kappelmann et al., 2018; Köhler-Forsberg et al., 2019; Wittenberg et al., 2020). To prioritise drug and patient selection for clinical trials, it is crucial to further understand immunological and clinical complexity of inflammation-symptom associations, which may allow shortlisting of promising immunotherapeutic drug targets and could highlight patients with a profile of inflammation-related depression.

Regarding immunological complexity, studies have reported various associations of serum inflammatory proteins with depression, including among others CRP, interleukin (IL)-6, IL-10, and tumour necrosis factor (TNFα)-(Goldsmith et al., 2016; Haapakoski et al., 2015; Köhler et al., 2017). Evidence from in-depth immunophenotyping further suggests that there may be distinct subgroups of inflammation-related depression as shown by immune cell count clustering and transcriptome analyses (Cattaneo et al., 2020; Lynall et al., 2020). These studies suggest that elevated serum levels of inflammatory markers are associated with depression, but associations of depression with genetic/lifetime predisposition to higher inflammatory markers has been studied less frequently and primarily for CRP (Badini et al., 2020; Kappelmann et al., 2021; Milaneschi et al., 2017b, 2016). Elevated serum levels of inflammatory markers also conflate tonic and phasic levels of inflammatory markers while genetic/lifetime predisposition to inflammatory markers specifically maps their tonic levels. This differentiation could be relevant as highlighted by research into tonic versus phasic dopamine levels (see Bilder et al., 2004), whereby tonic levels regulate the amplitude of the phasic response, which has unique consequences for downstream signalling. Lastly, inflammatory markers such as CRP are influenced by metabolic factors (Timpson et al., 2011), which may causally underlie some inflammation-symptom associations (Kappelmann et al., 2021), so a combined investigation of immuno-metabolic factors is needed to disentangle their etiological roles.

Regarding clinical complexity, most prior research has restricted its investigation of the inflammation-depression association to complexity on one side, that is focusing on multiple immune markers (e.g., cell counts/ serum cytokine levels) while studying a composite depression phenotype (Goldsmith et al., 2016; Haapakoski et al., 2015; Köhler et al., 2017) or focusing on multiple depressive symptoms or symptom groups in the context of a single inflammatory marker (mostly CRP) (Badini et al., 2020; Jokela et al., 2016; Köhler-Forsberg et al., 2017; Lamers et al., 2020, 2019; White et al., 2017). Among studies focusing on individual symptoms, results have highlighted associations of inflammatory markers with specific depressive symptoms of fatigue, changes in appetite, anhedonia, and suicidality (Badini et al., 2020; Chu et al., 2019; Jokela et al., 2016; Kappelmann et al., 2021; Köhler-Forsberg et al., 2017; Lamers et al., 2020, 2018; Milaneschi et al., 2017a; Simmons et al., 2018; White et al., 2017). However, most of these studies have considered associations of inflammatory markers with each depressive symptom in isolation (Chu et al., 2019; Jokela et al., 2016; Kappelmann et al., 2021; Köhler-Forsberg et al., 2017; Lamers et al., 2018; White et al., 2017). Although these prior approaches have led to important findings, they cannot address potential causal interactions between symptoms, thus conflate evidence for indirect and direct associations. For example, analyses of isolated symptoms could hypothetically provide evidence for associations of CRP with both fatigue and sleep problems even if CRP was only indirectly associated with fatigue *via* its effect on sleep problems. A network-based approach provides one means of disentangling such direct from indirect inflammation-symptom associations.

Network theory and related analysis techniques have recently been put forward to accommodate the symptomatic complexity of mental disorders (Borsboom, 2017). Network theory proposes putative causal interactions between symptoms (e.g., fatigue *causing* concentration problems *causing* low mood), which could result in self-reinforcing vicious symptom cycles triggering and maintaining mental disorders. Such associations have been investigated in an increasing amount of studies on psychological symptom networks (Contreras et al., 2019; Robinaugh et al., 2020). To accommodate etiological factors beyond symptoms, however, recent work has proposed an expansion of symptom networks to so-called ‘multi-plane’ networks, for instance also including genetic, metabolic, immunological, or environmental variables (Guloksuz et al., 2017). To our knowledge, so far, two studies have evaluated such multi-plane networks in the context of inflammation and depression by jointly analysing serum CRP (plus IL-6 & TNF-α in the study of Fried et al., 2019), BMI, and potential covariates with individual depressive symptoms (Fried et al., 2019; Moriarity et al., 2020a). The most consistently replicated findings between these two studies suggested unique associations of CRP with fatigue and changes in appetite. A third study has recently also provided evidence that the symptom structure itself was a function of CRP levels; that is, interconnections between symptoms were moderated by CRP (Moriarity et al., 2020b). All of these previous studies were based on serum markers for inflammatory proteins, however, reflective of acutely elevated inflammatory activity. Therefore, it remains unclear if inflammation-symptom associations generalise to genetic/lifetime predisposition to higher immuno-metabolic marker levels.

In the present study, we explored associations of polygenic risk scores (PRSs) for four major pro- and anti-inflammatory markers (i.e., CRP, IL-6, IL-10, & TNF-α) and Body Mass Index (BMI), as a metabolic marker, with individual depressive symptoms using a multi-sample, multi-plane network analysis approach. We evaluated associations in three large samples including the inpatient Munich Antidepressant Response Signature (MARS) study (n=1,058), the outpatient Sequenced Treatment Alternatives to Relieve Depression (STAR*D) study (n=1,143), and the general population UK Biobank cohort (n=110,010) (Hennings et al., 2009; Rush et al., 2004; Sudlow et al., 2015). This investigation aimed to contribute to the study of inflammation and depression by simultaneously addressing (i) combined immunological and symptom complexity (using network analysis), (ii) unclarity regarding the influence of genetic/lifetime predisposition to higher immuno-metabolic marker levels on depression (defining immuno-metabolic markers using PRSs), and (iii) issues of replicability and generalisability (testing associations in one large general population and two clinical samples).

## METHODS

An overview of the study design and analytic procedure is presented in Figure 1.

**Figure 1.**
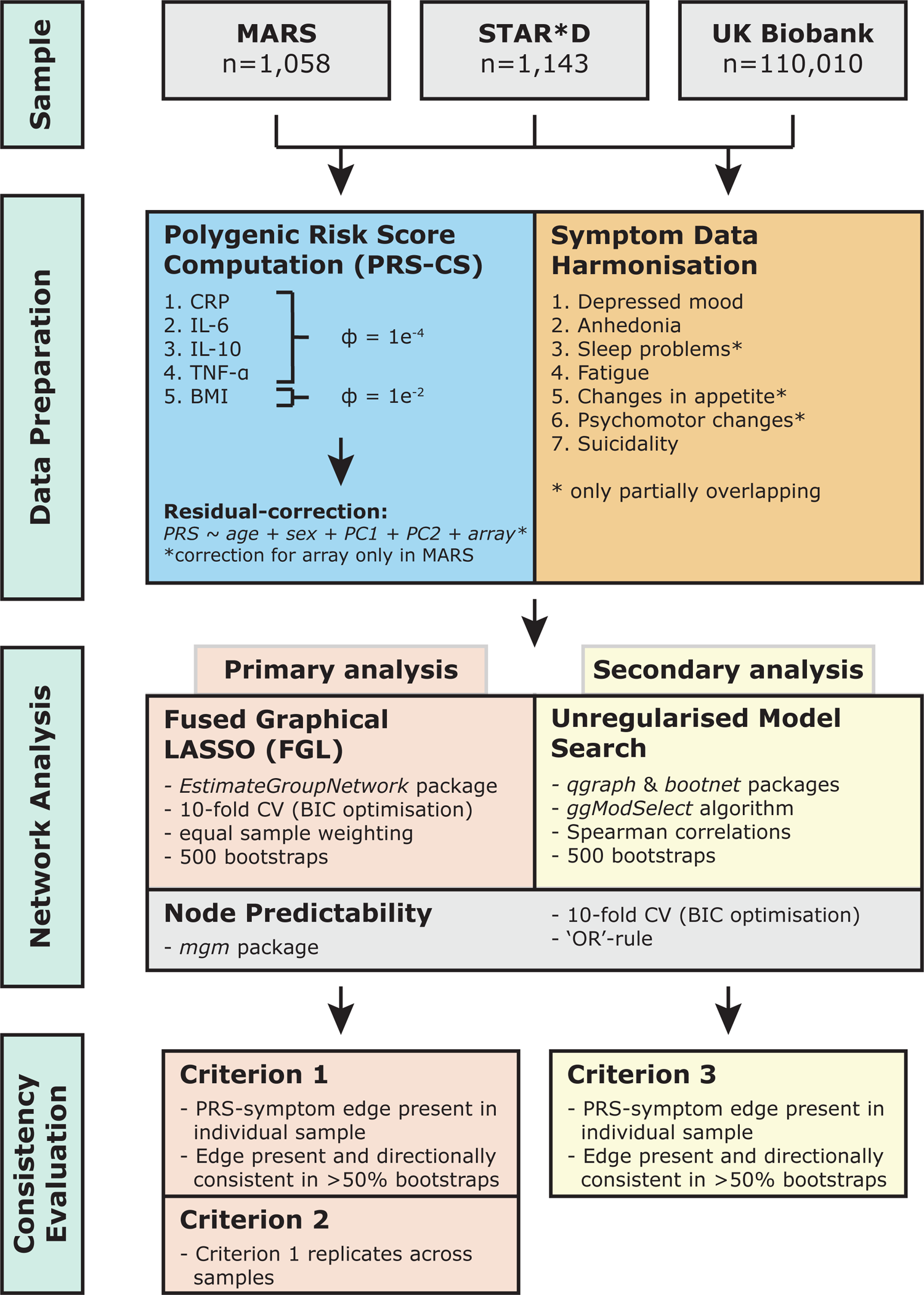
Study design and analysis pipeline. Legend: BIC=Bayesian information criterion; CV=cross-validation; PC=principal component (or multi-dimensional scaling component used for MARS & STAR*D); □ = PRS-CS tuning parameter.

### Study samples

The Munich Antidepressant Response Signature (MARS) study was a naturalistic, observational study of inpatients with major depressive disorder (MDD) or bipolar disorder conducted between 2000 and 2015 in three Southern German hospitals (Hennings et al., 2009). Based on an original sample of 1,411 patients, the present study included 1,058 patients of European decent with an ICD diagnosis of MDD (F32 and F33 codes) and genetic and depressive symptom data.

The STAR*D trial (identifier: NCT00021528) was a multisite, multistep, randomised controlled trial (RCT), conducted from 2000 to 2004, evaluating different treatment options and sequences for outpatients suffering from DSM-IV MDD without psychotic features (Rush et al., 2004).

Based on an original sample of 1,953 patients who took part in the STAR*D genetics study, the present study included 1,143 individuals of European decent with genetic and depressive symptom data.

The UK Biobank is a general population cohort including more than 500,000 individuals, recruited from 2006 to 2010, with genotyping and in-depth phenotyping information (Bycroft et al., 2018). More than 150,000 individuals from the initial sample took part in a follow-up mental health survey (Davis et al., 2020) and we included a subset of 110,010 individuals that were of European decent and had available genetic and depressive symptom data.

### Ethics approval and informed consent

MARS received local ethics approval from Ludwig Maximilians University Munich (Hennings et al., 2009). STAR*D received ethics approval from 14 participating institutional review boards, a National Coordinating Center, a Data Coordinating Center, and the Data Safety and Monitoring Board at the National Institute of Mental Health (Rush et al., 2006, 2004). The UK Biobank study received ethics approval from North West Centre for Research Ethics Committee and Human Tissue Authority research tissue bank (Bycroft et al., 2018); this project was approved under project no. 26999. All three studies collected informed consent from participants prior to study participation.

### Depressive symptom assessment

Depressive symptoms were assessed differently across the three samples. MARS and STAR*D studies used the observer-rated Hamilton Rating Scale for Depression (HAM-D) (Hamilton, 1986) while the UK Biobank study used the self-report Patient Heath Questionnaire (PHQ)-9 (Löwe et al., 2004). From these questionnaires, we selected seven depressive symptoms for joint analyses across samples. These symptoms included completely overlapping symptoms of depressed mood, anhedonia, fatigue, and suicidality, but also partially overlapping symptoms of sleep problems, changes in appetite, and psychomotor changes. Supplementary Table 1 provides an item-level overview of depressive symptoms and Supplementary Table 2 displays symptom coding, where this differed from original Likert scale ratings.

Regarding partially overlapping symptoms of sleep problems, changes in appetite, and psychomotor changes, the PHQ-9 only assesses information on conflated symptoms (e.g., insomnia and hypersomnia are conflated to sleep problems) while the HAM-D incorporates disaggregated symptoms. To harmonise these symptom data for retention in network analyses, we conflated HAM-D symptoms of psychomotor retardation and agitation to “psychomotor changes”. For sleep problems and changes in appetite (available in the PHQ-9), only insomnia and loss of appetite are available in the HAM-D, so we included both conflated and unidirectional symptoms in network analyses as previous studies have specifically highlighted associations of inflammation with these symptoms (Jokela et al., 2016; Milaneschi et al., 2017b). We reasoned that comparative appraisal of associations, for example with changes in appetite and loss of appetite, could give further indications on potential specificity of associations to symptom directions, as observed in previous reports (Kappelmann et al., 2021; Milaneschi et al., 2021b, 2021a).

We also note that we have not included items of “guilt or self-blame” from the respective studies in our analyses as we considered the item content of HAM-D and PHQ-9 too distinct.

Specifically, the HAM-D conflates feelings of guilt with delusions of guilt and death, thus moving towards psychotic symptomatology. Contrary to this, the PHQ-9 also includes “feelings of inadequacy” about oneself, which are not covered by the HAM-D item.

### Genotyping, quality control and imputation

We provide detailed information on genotyping, quality control and imputation procedures in the Supplementary Methods. Briefly, genotyping in the MARS study was conducted using three genotyping arrays across the recruitment period (see Supplementary Figure 1), the Illumina 610k (n=548), Illumina OmniExpress (n=284) and Illumina GSA (n=226) arrays. In STAR*D, genotyping was conducted using the Affymetrix Human Mapping 500K Array Set (n=979) and the Affymetrix Genome-Wide Human SNP Array 5.0 (n=969) that displayed a concordance of >99%; described in detail by Garriock *and colleagues* (2010). In the UK Biobank study, samples were genotyped on the UK BiLEVE Axiom Array or the Affymetrix UK Biobank Axiom Array (Bycroft et al., 2018). Following imputation in all samples, single nucleotide polymorphisms (SNPs) with info-metric>0.6, minor allele frequency (MAF)>1%, genotyping missingness<2%, and no deviation from Hardy-Weinberg Equilibrium (MARS & STAR*D: *P*>1e^-5^; UK Biobank: *P*>1e^-7^) were retained.

### Polygenic risk scores

#### Immuno-metabolic marker selection and GWAS data sources

PRSs for CRP, IL-6, IL-10, TNF-α, and BMI were computed based on available summary statistics from genome-wide association studies (GWAS; Ahola-Olli et al., 2017; Ligthart et al., 2018; Locke et al., 2015). These inflammatory markers were selected, because (i) they showed robust differences in case-control studies; (ii) CRP, IL-6, and TNF-α have been the most frequently investigated inflammatory markers overall in the context of depression; and (iii) IL-10 was the most frequently studied anti-inflammatory cytokine, so could be informative on direction of associations between depressive symptoms and innate immune activity (Köhler et al., 2017; Osimo et al., 2019). BMI was selected as the most frequently investigated metabolic marker.

GWAS data for CRP were obtained from a large GWAS of 88 studies including 204,402 individuals of European decent (Ligthart et al., 2018). GWAS data for IL-6, IL-10, and TNF-α were obtained from a GWAS of 8,293 Finns (Ahola-Olli et al., 2017); of note, Finns have Siberian ancestry (Lamnidis et al., 2018), which leads to a divergence from European ancestry of our analytic samples. GWAS data for BMI were obtained from the Genetic Investigation of Anthropometric Traits (GIANT) consortium that included up to 322,154 individuals of European decent (Locke et al., 2015).

#### PRS computation

PRSs can be computed by summing the GWAS association estimates of risk alleles for each individual. Classically, this summation is done using an approach termed “clumping and thresholding” (C+T), which first reduces summary statistics to independent SNPs and then applies one or multiple thresholds (usually based on P-values) to restrict summation to SNPs with high evidence for associations with phenotypes (Choi et al., 2020). As the optimal threshold for the C+T approach is unknown and should ideally be estimated in a separate dataset with available phenotype data, we computed PRSs using the Bayesian regression and continuous shrinkage priors (PRS-CS) approach, which has been shown to perform similar to or outperform other PRS computation approaches such as C+T (Ge et al., 2019; Ni et al., 2020).

PRS-CS takes a linkage disequilibrium (LD) reference panel into account (we used European ancestry data from 1000 Genomes Project phase 3 samples) to update SNP effect sizes in a blocked fashion, thus providing accurate LD adjustment. We pre-specified the global shrinkage parameter □ using suggested defaults for less polygenic (□ =1e^-4^) and more polygenic (□ =1e^-2^) phenotypes as □ =1e^-4^ for CRP, IL-6, IL-10, and TNF-α, and as □ =1e^-2^ for BMI; see details in Supplementary Methods. Following PRS computation in individual samples, polygenic scores were corrected for age, sex, and the first two genotyping principal or multidimensional scaling (MDS) components using linear regression; two genotyping principal or MDS components were selected as visual inspection of component inter-correlations did not suggest evidence for population stratification. Genotyping MDS components were computed based on raw Hamming-distances in MARS and STAR*D, and using principal component analysis on high-quality, unrelated individuals in the UK Biobank sample (Bycroft et al., 2018). PRSs in MARS were additionally corrected for the genotyping array. Following computation, higher PRSs reflect higher genetic predisposition to respective immuno-metabolic phenotype levels.

#### PRS evaluation

In Supplementary Table 3, we provide the number of SNPs included in PRS computation in each sample, which was approximately around one million SNPs for each phenotype-sample combination. The proportion of SNP overlap between samples (for the same phenotype) was >0.89 suggesting that mostly overlapping SNPs contributed to PRSs (Supplementary Table 4). Taking these overlapping SNP sets, correlations between the posterior SNP effect sizes between samples were large for CRP (Pearson’s *r* range: 0.69-0.76) and BMI (Pearson’s *r* range: 0.79-0.80) and relatively smaller for IL-6, IL-10, and TNF-α (Pearson’s *r* range: 0.41-0.46; see Supplementary Table 5). This suggests polygenic risk was quantified more similarly across samples for CRP and BMI as compared to IL-6, IL-10, and TNF-α.

We quantified the impact that pre-specification of the hyperparameter LJ had on resulting PRSs, which was likely small (Supplementary Table 6). Specifically, PRSs with pre-specified exhibited large correlations with PRSs based on automatic learning of from GWAS summary data (termed PRS-CS-auto in the literature; Pearson’s *r* range: 0.82-0.98). Furthermore, moderate-to-large correlations remained to PRSs based on extreme grid search boundary values of (Pearson’s *r* range: 0.47-0.93).

Since MARS utilised three different genotyping arrays, we verified that our approach of combining data from these arrays into one sample was justified before proceeding with the main analysis (see Supplementary Methods and Supplementary Figures 2 and 3).

**Figure 2.**
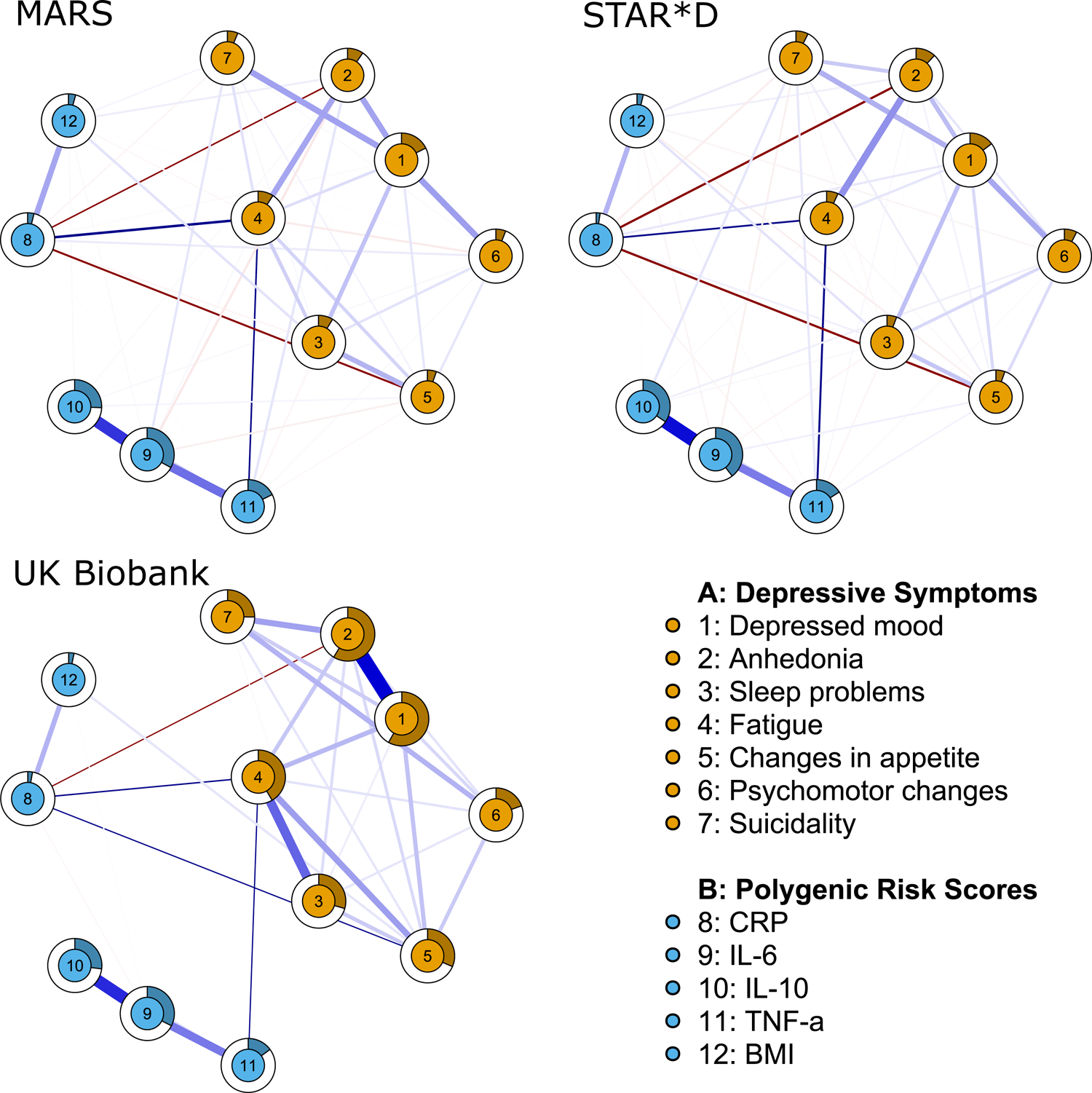
Estimated FGL networks across samples. Legend: Networks are visualised with the *qgraph* package. Blue lines indicate positive and red lines negative associations, respectively, with larger associations displayed with thicker lines. Circles around nodes display node predictability, which can be interpreted similar to explained variance. Maximum size of edge associations is 0.55. As the primary focus of this investigation was to identify consistent PRS-symptom associations, we manually unfaded edges between PRSs and symptoms if these edges met quality criteria 1 and 2 (see Table 2). Changes in appetite and sleep problems are measured as composite symptoms in UK Biobank, but as loss of appetite and insomnia in MARS and STAR*D samples.

**Figure 3.**
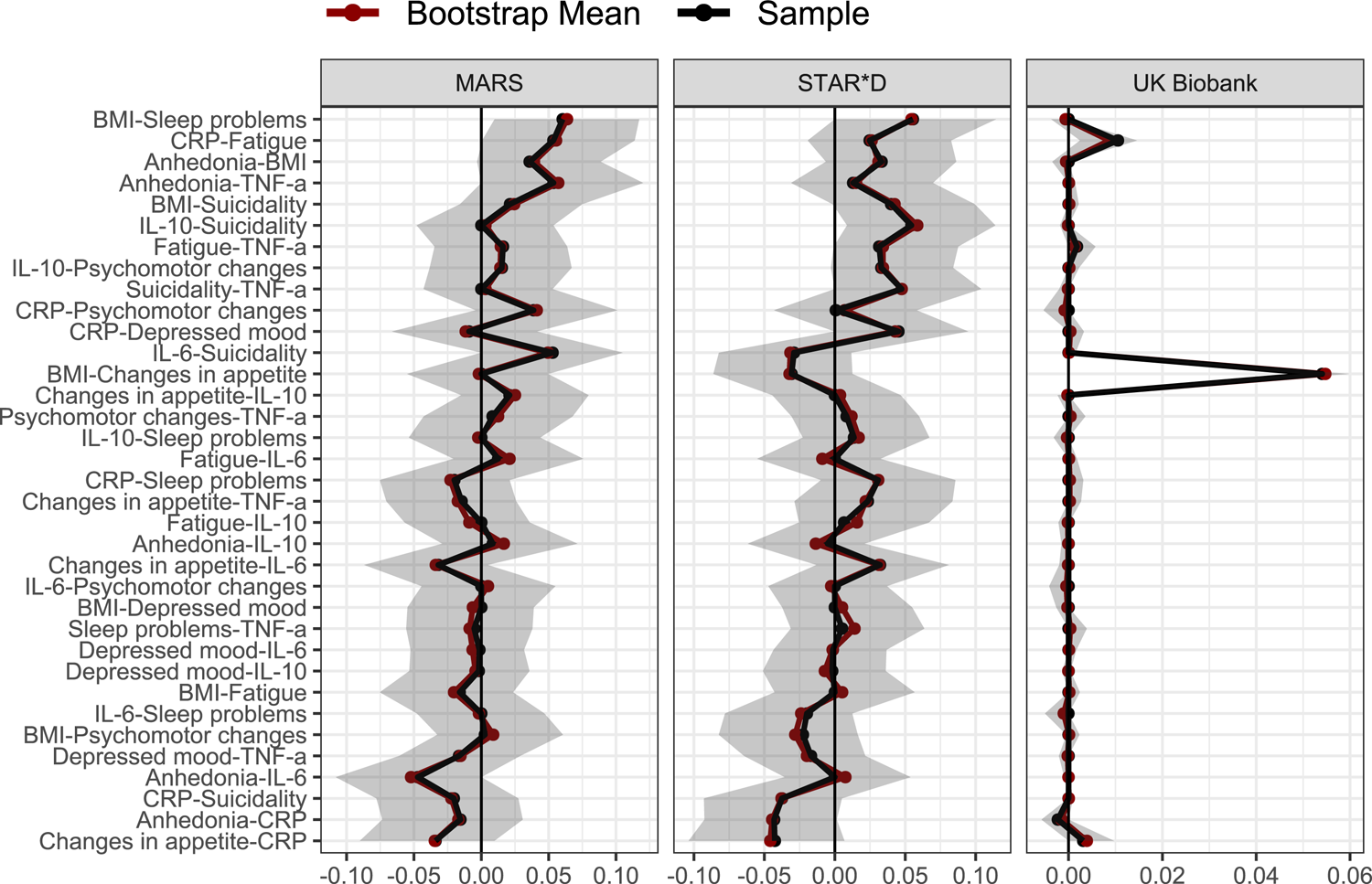
Bootstrapped 95% quantile intervals of PRS-symptom edges using FGL estimation. Legend: Bootstrapped 95% quantile intervals (i.e., 95% of the distribution of raw bootstrapped edge estimates) are highlighted as shaded area for each edge. Black points indicate the raw FGL sample estimate while red points indicate the raw bootstrapped mean estimate. Edges are indicated on the y-axis and sorted by mean edge weight across samples in descending order.

### Network analysis

#### Estimation

Network analysis was conducted using *R* software (version 4.0.3; R Core Team, 2017). In network analysis, unique associations between variables reflect partial correlations and are termed ‘edges’. Variables in the network are referred to as ‘nodes’.

Network models can be broadly categorised into regularized and unregularized models, that have distinct advantages and disadvantages. Regularised models apply penalties that shrink edges towards zero. This has the advantage that it results in sparser and more parsimonious network models as small edges can be set exactly to zero. Contrary to this, non-regularized models do not apply such a penalty while still controlling the false positive rate and recent studies have suggested that unregularized models perform better in estimating psychological symptom networks and multi-plane immunopsychiatric networks than regularized network models (Moriarity et al., 2020a; Williams et al., 2019). A disadvantage of unregularized models, however, is that they are currently only suitable for network estimation of individual samples/datasets. Contrary to this, regularised models have recently been adapted for application in multi-sample contexts using so-called fused graphical LASSO (FGL) estimation. FGL estimation allows synthesising data across multiple samples, which increases statistical power.

Based on these respective advantages and disadvantages, we have decided to use a regularized network model as primary analysis, which maximises statistical power due to the multi-sample design of our study. As unregularized models are preferable for estimation of individual samples and may be better suited to retrieve multi-plane edges, however, we also apply unregularized network estimation as secondary analysis.

In primary analyses, networks were estimated using FGL estimation as implemented in the *EstimateGroupNetwork* package (version 0.2.2; Costantini et al., 2020, 2019; Danaher et al., 2014). FGL estimation relies on the two tuning parameters λ_1_, which penalizes network density, and λ_2_, which penalizes edge differences across samples. Values for these tuning parameters were selected using 10-fold cross-validation to optimise the Bayesian Information Criterion (BIC). As recommended, we set weights for the importance of each sample as ‘equal’ to ascertain that a single sample would not dominate estimation (Danaher et al., 2014).

As secondary analysis, we estimated unregularized networks for each sample individually using the gaussian graphical stepwise model selection (“ggModSelect”) algorithm implemented in the *qgraph* package (version 1.6.5; Epskamp et al., 2012) based on Spearman correlations and starting from an empty model. Throughout results, we refer to this estimation strategy as “unregularized model search” or “model search” for simplification.

#### Node predictability

We also estimated node predictability, which describes the amount of variance in a node that is explained by all other nodes in the network, so can be interpreted akin to R^2^ (Haslbeck and Fried, 2017). Node predictability cannot be inferred from FGL or model search networks as it requires a node-wise estimation approach. Therefore, we used a mixed graphical model as a third estimation strategy as implemented in the *mgm* package (version 1.2-10), selecting tuning parameter λ based on BIC optimisation in 10-fold cross-validation (Haslbeck and Waldorp, 2020). Of note, this model was only used to infer node predictability, which provides additional information on network density and sample comparability. However, we do not report any individual edge estimates based on this model as FGL estimation and unregularized model search are better suited for our study aims.

#### Visualisation

Networks were visualised with the *qgraph* package using an average layout estimated with the Fruchterman-Reingold algorithm for the FGL networks. This algorithm places nodes close to each other that are connected by large edges (Epskamp et al., 2012). While this simplifies network appraisal, it is important to note that nodes and edges should not be interpreted based on their relative position within the network, which can be unstable.

#### Stability

To evaluate stability of estimated networks, we assessed accuracy of edge estimates using bootstrapping strategies. Specifically, for FGL networks 500 bootstrapped samples with replacement were drawn, and FGL networks re-estimated, using the implementation in the *EstimateGroupNetwork* package (Costantini et al., 2020). For unregularized model search estimation, the same procedure was applied using non-parametric bootstrapping procedures implemented in the *bootnet* package (version 1.4.3; Epskamp et al., 2018).

#### Interpretation

We interpreted estimated networks based on the presence, stability, and replicability of edges as defined using three consistency criteria. First, we tested if edges were nonzero in FGL networks as well as nonzero and directionally consistent in >50% of bootstrapped analyses (consistency criterion 1) akin to a previous PRS-symptom network study in psychosis by Isvoranu *and colleagues* (2020). Second, we tested if edges between PRSs and symptoms replicated (according to criterion 1) across FGL networks of the three samples (consistency criterion 2). Third, we tested if edges were present in secondary analyses using unregularized model search estimation, again confirmed in >50% of bootstrapped estimations exhibiting directionally consistent estimates (consistency criterion 3).

### Availability of data and materials

Data from original studies is not openly available, but can be requested; see details in Supplementary Table 7. GWAS summary data for IL-6, IL-10, and TNF-α is openly available from the original publication by Ahola-Olli *and colleagues* (2017), for BMI from the GIANT consortium, and can be requested for CRP from the CHARGE inflammation working group. We provide analysis scripts and estimated network matrices (including bootstrapped network matrices) on the Open Science Platform (OSF) under https://osf.io/q4vw9/.

## RESULTS

Baseline characteristics of study populations are displayed in Table 1.

**Table 1.** Baseline characteristics of MARS, STAR*D, and UK Biobank samples

|  | **MARS** | **STAR\*D** | **UK Biobank** |
| --- | --- | --- | --- |
| N | 1,058 | 1,143 | 110,010 |
| Sex |  |  |  |
| Women, N (%) | 563 (53.2%) | 676 (59.1%) | 61,212 (55.6%) |
| Men, N (%) | 495 (46.8%) | 467 (40.9%) | 48,798 (44.4%) |
| Age in years |  |  |  |
| Mean (SD) | 47.8 (14.4) | 43.2 (13.6) | 56.2 (7.7) |
| Range | 18-87 | 18-75 | 39-72 |
| Study location | Germany | United States | United Kingdom |
| Study population | MDD inpatients | MDD outpatients | General population |
| CRP PRS |  |  |  |
| Mean (SD) | -0.01 (1.00) | 0.00 (1.00) | -0.01 (1.00) |
| Range | -3.14-3.15 | -3.21-2.94 | -4.58-4.21 |
| IL-6 PRS |  |  |  |
| Mean (SD) | 0.00 (1.00) | 0.00 (1.00) | 0.00 (1.00) |
| Range | -3.33-3.34 | -3.41-3.94 | -4.07-3.92 |
| IL-10 PRS |  |  |  |
| Mean (SD) | 0.00 (1.00) | 0.00 (1.00) | 0.00 (1.00) |
| Range | -3.11-3.24 | -3.28-3.13 | -4.35-4.60 |
| TNF-α PRS |  |  |  |
| Mean (SD) | 0.00 (1.00) | 0.00 (1.00) | 0.00 (1.00) |
| Range | -3.61-3.67 | -3.35-3.52 | -4.17-4.51 |
| BMI PRS |  |  |  |
| Mean (SD) | -0.02 (1.01) | 0.00 (1.00) | -0.04 (1.00) |
| Range | -3.02-4.06 | -3.04-3.20 | -4.53-3.96 |
| PHQ-9 sum-score |  |  |  |
| Mean (SD) | - | - | 2.7 (3.6) |
| Range | - | - | 0-27 |
| Missing, N (%) | - | - | 351 (0.3) |
| HAM-D sum-score |  |  |  |
| Mean (SD) | 23.8 (5.9) | 22.4 (4.9) | - |
| Range | 5-42 | 13-38 | - |
| Missing, N (%) | 6 (0.6%) | 0 (0%) | - |
| Depressed mood |  |  |  |
| Mean (SD) | 3.05 (0.88) | 2.59 (0.77) | 0.23 (0.56) |
| Range | 0-4 | 0-4 | 0-3 |
| Anhedonia |  |  |  |
| Mean (SD) | 3.63 (0.68) | 2.54 (0.76) | 0.26 (0.56) |
| Range | 0-4 | 0-4 | 0-3 |
| Sleep problems |  |  |  |
| Mean (SD) | 3.46 (2.00) | 3.42 (1.77) | 0.71 (0.99) |
| Range | 0-6 | 0-6 | 0-3 |
| Fatigue |  |  |  |
| Mean (SD) | 1.49 (0.68) | 1.68 (0.55) | 0.66 (0.81) |
| Range | 0-2 | 0-2 | 0-3 |
| Changes in appetite |  |  |  |
| Mean (SD) | 0.77 (0.66) | 0.68 (0.80) | 0.25 (0.62) |
| Range | 0-2 | 0-2 | 0-3 |
| Psychomotor changes |  |  |  |
| Mean (SD) | 1.44 (0.96) | 1.42 (0.72) | 0.07 (0.34) |
| Range | 0-4 | 0-4 | 0-3 |
| Suicidality |  |  |  |
| Mean (SD) | 1.35 (1.15) | 0.94 (0.85) | 0.05 (0.28) |
| Range | 0-4 | 0-4 | 0-3 |

*Note:* MDD=Major Depressive Disorder, SD=Standard deviation, PHQ-9=Patient Health Questionnaire-9, HAM-D=Hamilton Rating Scale for Depression.

### Network analysis

We conducted network analyses of five immuno-metabolic PRSs (CRP, IL-6, IL-10, TNF-α & BMI) and seven depressive symptoms using two estimation techniques (FGL & unregularized model search estimation) in three samples (MARS, STAR*D & UK Biobank). Bootstrap analyses were conducted to assess stability of networks and node predictability estimated using a mixed graphical model. We defined three consistency criteria to assess robustness and replicability of our results across estimation techniques, bootstrapping, and samples. Focus of this network investigation were unique associations (termed edges in network analysis) between PRSs and symptoms, which are summarised in Table 2.

**Table 2.** PRS-symptom edge consistency criteria (C) across network analyses

| PRS-symptom edges | MARS |  | STAR*D |  | UK Biobank |  | FGL consistency (C2) |
| --- | --- | --- | --- | --- | --- | --- | --- |
|  | FGL (C1) | Model search (C3) | FGL (C1) | Model search (C3) | FGL (C1) | Model search (C3) |  |
| **CRP** |  |  |  |  |  |  |  |
| Anhedonia | -0.016 (67%) |  | -0.043 (95%) |  | -0.002 (60%) |  | Yes |
| Depressed mood | -0.009 (57%) |  | 0.045 (94%) |  |  |  |  |
| Sleep problems* | -0.02 (75%) |  | 0.031 (84%) |  |  |  |  |
| Fatigue | 0.053 (98%) |  | 0.025 (79%) |  | 0.011 (100%) |  | Yes |
| Changes in appetite* | -0.034 (89%) |  | -0.043 (94%) |  | 0.003 (91%) | 0.013 (73%) | Yes |
| Psychomotor changes | 0.039 (91%) |  | 0.001 (53%) |  |  |  |  |
| Suicidality | -0.02 (74%) |  | -0.038 (88%) |  |  |  |  |
| **IL-6** |  |  |  |  |  |  |  |
| Anhedonia | -0.047 (97%) |  |  |  |  |  |  |
| Depressed mood | -0.001 (52%) |  |  |  |  |  |  |
| Sleep problems* |  |  | -0.02 (82%) |  |  |  |  |
| Fatigue | 0.012 (72%) |  |  |  |  |  |  |
| Changes in appetite* | -0.032 (87%) |  | 0.032 (85%) |  |  |  |  |
| Psychomotor changes |  |  |  |  |  |  |  |
| Suicidality | 0.053 (95%) |  | -0.029 (83%) |  |  |  |  |
| **IL-10** |  |  |  |  |  |  |  |
| Anhedonia | 0.008 (67%) |  | -0.005 (66%) |  |  |  |  |
| Depressed mood |  |  | -0.002 (52%) |  |  |  |  |
| Sleep problems* |  |  | 0.014 (72%) |  |  |  |  |
| Fatigue |  |  | 0.007 (66%) |  |  |  |  |
| Changes in appetite* | 0.021 (80%) |  |  |  |  |  |  |
| Psychomotor changes | 0.015 (66%) |  | 0.033 (90%) |  |  |  |  |
| Suicidality |  |  | 0.054 (99%) |  |  |  |  |
| **TNF-α** |  |  |  |  |  |  |  |
| Anhedonia | 0.054 (96%) |  | 0.013 (66%) |  |  |  |  |
| Depressed mood | -0.017 (66%) |  | -0.017 (75%) |  |  |  |  |
| Sleep problems* | -0.005 (57%) |  | 0.005 (68%) |  |  |  |  |
| Fatigue | 0.016 (65%) |  | 0.032 (91%) |  | 0.002 (58%) |  | Yes |
| Changes in appetite* | -0.015 (70%) |  | 0.023 (75%) |  |  |  |  |
|  | FGL (C1) | Model search (C3) | FGL (C1) | Model search (C3) | FGL (C1) | Model search (C3) |  |
| Psychomotor changes | 0.008 (63%) |  | 0.008 (59%) |  |  |  |  |
| Suicidality |  |  | 0.047 (94%) |  |  |  |  |
| **BMI** |  |  |  |  |  |  |  |
| Anhedonia | 0.036 (91%) |  | 0.033 (86%) |  |  | -0.010 (63%) |  |
| Depressed mood |  |  |  |  |  |  |  |
| Sleep problems* | 0.06 (99%) |  | 0.055 (97%) |  |  |  |  |
| Fatigue | -0.016 (74%) |  |  |  |  |  |  |
| Changes in appetite* |  |  | -0.031 (87%) |  | 0.054 (100%) | 0.066 (100%) |  |
| Psychomotor changes | 0.003 (55%) |  | -0.023 (83%) |  |  |  |  |
| Suicidality | 0.021 (75%) |  | 0.04 (91%) |  |  |  |  |

*Note:* Cell values reflect edge weights (i.e., partial correlation coefficients) and the percentage of 500 bootstrap estimations that edges were present. Estimates are restricted to those edges, for which >50% of bootstrapped samples were non-zero and directionally consistent (i.e., criteria 1 & 3). \* Changes in appetite and sleep problems are measured as composite symptoms in UK Biobank, but as loss of appetite and insomnia in MARS and STAR\*D samples.

### Fused Graphical LASSO (FGL) estimation suggests four consistent PRS-symptom edges according to criteria 1 & 2

Using FGL estimation, we obtained networks that are visualised in Figure 2. PRS-symptom edge bootstrapping results are displayed in Figure 3 with PRS-PRS and symptom-symptom edge bootstrapping results shown in Supplementary Figures 4 and 5.

As expected, nodes within the same plane displayed relatively stronger within-plane (i.e., symptom-symptom & PRS-PRS) than between-plane (i.e., PRS-symptom) associations. Among PRSs, CRP displayed associations with BMI (edge weight range across samples: 0.16-0.19) while IL-6, IL-10, and TNF-α (based on the same GWAS) were associated with each other (edge weight range across samples: 0.08-0.52). Associations of BMI and CRP with IL-6, IL-10, and

TNF-α were largely absent or very small (edge weight range across samples: −0.02-0.01). Among symptoms, the largest associations were present between the core symptoms depressed mood and anhedonia (edge weight range across samples: 0.14-0.55), which is to-be-expected in clinical samples where these symptoms form the basis of the MDD diagnosis. Supplementary Figure 4 also illustrates interesting edge differences between samples that are likely arising from the diverging symptom definitions in individual samples. For instance, edges of fatigue with changes in appetite (edge weight=0.21) and sleep problems (edge weight=0.33) were relatively larger in the UK Biobank, assessing composite symptoms of changes in appetite and sleep problems, but substantially smaller in MARS (fatigue-changes in appetite: edge weight=0.09; fatigue-sleep problems: edge weight=0.10) and STAR*D (fatigue-changes in appetite: edge weight=-0.01; fatigue-sleep problems: edge weight=-0.01), assessing loss of appetite and insomnia.

Regarding PRS-symptom edges, FGL estimation surprisingly resulted in a much larger number of PRS-symptom edges in MARS and STAR*D samples compared to the UK Biobank sample with 28 (MARS), 29 (STAR*D), and 6 (UK Biobank) nonzero PRS-symptom edges. 26 (MARS), 28 (STAR*D), and 5 (UK Biobank) of these edges fulfilled criterion 1 (nonzero edges are nonzero and directionally consistent in >50% of bootstraps). Although the difference between samples could have resulted from network differences of clinical versus general population-based samples, it may also reflect some degree of inconsistency or even noise as edge estimates often exhibited unstable directions of association in clinical samples (see Table 2).

Applying consistency criterion 2 (consistency of results across samples), we observed replicable edges of the CRP PRS with anhedonia (negative edge weight), changes in appetite, and fatigue and of the TNF-α PRS with fatigue; these edges were manually unfaded in Figure 2. It is important to note that the edge between the CRP PRS and changes in appetite has a diverging valence in individual samples; in MARS and STAR*D (assessing loss of appetite) the edge weight was negative and in the UK Biobank study (assessing changes in appetite) the edge weight was positive.

### Unregularized model search estimation suggests three consistent PRS-symptom edges according to criterion 3

Using unregularized model search estimation, we again observed networks with relatively larger within-plane (i.e., PRS-PRS & symptom-symptom) than between-plane (i.e., PRS-symptom) edges. Networks were comparable to FGL estimation, but generally sparser than those using FGL estimation; see network graphs in Supplementary Figure 6 and bootstrapping results in Supplementary Figures 7-9.

Regarding PRS-symptom edges, only three edges were estimated, which were all observed in the UK Biobank sample and fulfilled consistency criterion 3 (nonzero edges are also nonzero and directionally consistent in >50% of bootstraps); these edges have been manually unfaded in Supplementary Figure 6. The specific PRS-symptom edges were between the BMI PRS and changes in appetite and anhedonia and between the CRP PRS and changes in appetite.

Comparing these edges to FGL estimation, the edge of the CRP PRS with changes in appetite replicated one of the edges fulfilling consistency criteria 1 and 2 while the two edges observed for the BMI PRS were only fulfilling consistency criterion 1 (presence in FGL estimation and >50% of bootstraps). Moreover, the BMI PRS association with anhedonia was negative using unregularized model search estimation, but positive using FGL estimation.

### Node predictability

Average node predictability was similar across samples for PRS nodes with 16% (UK Biobank), 17% (MARS), and 19% (STAR*D) of variance explained by all other nodes in the network.

Contrary to this, average node predictability for symptom nodes differed with 38% of variance explained by all other nodes in the UK Biobank sample and only 9% in both clinical samples. These findings highlight differences in network density of symptoms in the UK Biobank (using the PHQ-9) and clinical samples (using the HAM-D).

## DISCUSSION

The present study investigated associations of PRSs for immuno-metabolic markers with depressive symptoms using a multi-plane, multi-sample network analysis approach. Based on three consistency criteria emphasising robustness and replicability of network analysis results across statistical bootstraps, samples, and estimation methods, we observed a unique association between the CRP PRS and changes in appetite that met all three consistency criteria. In addition to this association, we observed five additional PRS-symptom associations that met two consistency criteria. These included edges of the CRP PRS with anhedonia (negative association) and fatigue, the TNF-α PRS with fatigue, and the BMI PRS with anhedonia and changes in appetite. However, the BMI PRS-anhedonia association switched association direction depending on the estimation method, so may not be fully consistent despite fulfilling our consistency criteria. Due to the novelty of our analysis approach, we highlight several methodological considerations below, which we hope provides a helpful framework to the discussion of our findings afterwards.

### Methodological challenges and opportunities

Combining PRSs with psychological symptom networks is a relatively recent extension of network analysis and, to our knowledge, has only been applied in one previous investigation incorporating a schizophrenia PRS into a psychotic symptom network (Isvoranu et al., 2020). Therefore, it is important to emphasise the unique challenges and opportunities of this approach.

First, as noted by Isvoranu *and colleagues* (2020), statistical power is potentially the greatest challenge of PRS-symptom network analysis. Network analysis itself requires relatively large sample sizes for psychological symptom networks (Epskamp et al., 2018; Fried and Cramer, 2017), which should be in the hundreds or thousands depending on the number of nodes in the network. Inclusion of PRSs into psychological symptom networks, and especially of potential pathomechanistic (e.g., inflammatory) rather than main illness (e.g., depression/schizophrenia) scores into these networks, aggravates the sample size requirements for network analysis as PRSs only explain a fraction of variance in the heritable component of their target phenotypes (Choi et al., 2020; Wray et al., 2020).

Second, and because PRSs only measure a fraction of variance in their target phenotype, unique associations observed in network analyses are inevitably smaller than actual target phenotype-symptom associations. Taking this study as an example, absolute sizes of CRP PRS-symptom associations were 5- to 10-fold smaller than those from a prior network investigation using serum CRP concentrations by Moriarity *and colleagues* (2020a). Therefore, PRS-symptom associations are unlikely to give meaningful insights into size of association with the target phenotype, but should, in our opinion, be interpreted based on robust presence/absence of specific associations.

Third, the large statistical power requirements and difficulty quantifying such power for a given study may lead to biased result interpretations. Absence of PRS-symptom associations could be interpreted as false negatives while presence of association may be interpreted as true positives. Such divergence in interpretation necessarily biases the literature towards hypothesis confirmation. Consequently, any associations observed in PRS-symptom network analyses should be followed up by- and interpreted in line with-evidence from other studies, thus adhering to the recommended triangulation of evidence approach (Lawlor et al., 2017; Ohlsson and Kendler, 2019).

Despite these challenges, PRS-symptom networks also provide multiple opportunities. First, PRSs reflect estimates of genetic liability to phenotype expression, so can give an indication on the influence of lifelong predisposition to higher phenotype levels on the symptom level. In this way, PRS-symptom associations also provide an indication regarding temporality of association, which Bradford-Hill defined as one of the viewpoints for causality (Bradford Hill, 1965). It is important to note, however, that evidence for a unidirectional temporal association does not preclude bi-directionality. Moreover, PRSs combine information from a multitude of genetic variants (in our case from ∼1 million SNPs) that are not restricted to functional SNPs, can include false positive associations (i.e., noise), and can also tag information of pleiotropic environmental confounding factors. Therefore, causal inferences should rely on separate evidence from clinical trials and/or more focused genetic approaches such as Mendelian Randomisation studies (Lawlor et al., 2008).

Second, the PRS-symptom network analysis approach allows the concurrent investigation of multiple immuno-metabolic markers with multiple symptoms. Thereby, immunological and clinical complexity is addressed concurrently, which is an advantage to previous investigations. Furthermore, network analyses usually estimate partial/unique associations, so any emerging associations could suggest direct causal paths from PRS phenotypes to individual symptoms, so may pinpoint so-called ‘bridge symptoms’ that act as etiological docking sites of risk effects on the symptom plane.

Third, large-scale population-based or patient cohort studies, commonly used in network analysis, often do not have detailed immunophenotyping data available. If at all, studies mostly have data available for serum CRP, but rarely for more specific cytokines. Conversely, the advent of large GWAS investigations has produced a substantial amount of large cohort databases with in-depth genotyping and phenotyping information. Combining such databases with GWAS summary statistics from more focused investigations, such as on individual cytokines (Ahola-Olli et al., 2017), enables the investigation of a diverse range of immunopsychiatric research questions.

Fourth, PRS-symptom networks could be extended, for instance, by adding serum inflammatory markers to these networks, which could provide additional insights into associations between genetic/lifelong predisposition to, and acute levels of, immuno-metabolic markers with individual symptoms.

### Associations of immuno-metabolic markers with depressive symptoms

Network analysis results showed consistent associations of the CRP PRS with changes in appetite, which was the only association that fulfilled all of our quality criteria. The BMI PRS showed similar associations with changes in appetite, but only fulfilled two quality criteria.

Importantly, both of these associations were positive in the UK Biobank sample, which assessed changes in appetite, and negative in MARS and STAR*D samples, which assessed loss of appetite. Previous studies reporting results from cross-sectional, longitudinal, genetic correlation, PRS, and Mendelian randomisation analyses have also consistently reported associations of CRP/BMI with changes in appetite (Fried et al., 2019; Jokela et al., 2016; Kappelmann et al., 2021; Moriarity et al., 2020a). Importantly, whenever studies disaggregated appetite symptoms into decreased versus increased appetite, associations of CRP/BMI were specific to increased appetite (Lamers et al., 2018; Milaneschi et al., 2021b, 2021a, 2017b; Pistis et al., 2021; Simmons et al., 2018). In light of these findings, our results provide indirect support for an immune-metabolic contribution to increased appetite specifically.

In addition to these PRS associations with changes in appetite, we also observed associations of higher CRP PRS with lower anhedonia and greater fatigue and of higher TNF-α PRS with greater fatigue. Fatigue in particular has long been considered to have a neuroimmune basis (Dantzer et al., 2014), is common across other medical illnesses characterised by chronic inflammation, and has been reliably associated with inflammatory markers in previous studies including two network investigations (Fried et al., 2019; Jokela et al., 2016; Lamers et al., 2020; Moriarity et al., 2020a; van Eeden et al., 2020; White et al., 2017). While there have also been some studies suggesting associations of inflammatory markers with anhedonia (Köhler-Forsberg et al., 2017; van Eeden et al., 2020), it is important to note that associations of the CRP PRS with anhedonia observed in the present report were negative, so do not offer straightforward replication of these findings. Nonetheless, we have recently shown in Mendelian randomisation analyses that BMI could be a potential causal factor for both fatigue and anhedonia (Kappelmann et al., 2021), so continued investigation of these symptoms is warranted.

Together, our findings add to the notion of an immuno-metabolic subtype of depression characterised by neurovegetative symptoms of changes in appetite and fatigue (Dantzer et al., 2008; Milaneschi et al., 2020). We also expand upon previous work by showing that genetic/lifetime predisposition to higher inflammation and metabolic dysregulation increases risk for depression and, based on network analysis results, these etiological factors may specifically confer their risk on the broader depression syndrome through symptoms such as changes in appetite and fatigue. These results can inform the design of clinical trials of anti-inflammatory approaches and metabolic interventions by specifically selecting patients with an atypical, neurovegetative symptom presentation. As clinical trials for immune-modulating drugs are currently still characterised by relatively small sample sizes (Husain et al., 2020; Khandaker et al., 2018; McIntyre et al., 2019; Nettis et al., 2021; Raison et al., 2013), it may be worthwhile to pilot new interventions with neurovegetative symptoms/phenotypes as outcome variables. This might increase statistical power and sensitivity to detect effects for these proof-of-concept trials and could then be followed up by larger trials testing broader clinical efficacy measures.

### Strengths and limitations

Strength of this study include availability of large general population-based and patient samples (maximising replicability and generalisability), polygenic definition of immuno-metabolic risk variables (indexing lifetime predisposition to higher immuno-metabolic marker levels), and application of network analysis (addressing immunological and clinical complexity concurrently). We have addressed some of the more general limitations of combined PRS-symptom network analysis above, but there are three more specific limitations that warrant mentioning.

First, data used in the current study included inpatients, outpatients, and individuals from the general population and was based on different scales to measure depressive symptoms.

Depressive symptom structure varies between acutely ill patients versus those in remission (van Borkulo et al., 2015), which may have influenced PRS-symptom associations. Moreover, two of the seven symptoms used in the present report only overlap partially; the UK Biobank study includes conflated items on sleep problems and changes in appetite while MARS and STAR*D include items on insomnia and loss of appetite, respectively. This difference may explain some of the inconsistencies observed in the current report such as the diverging valence of edge estimates between CRP and changes in appetite. However, this may have also reduced statistical power to detect associations. Study questionnaires also differed regarding the method of assessment as the HAM-D is observer-rated and the PHQ-9 self-reported. By definition, inflammation-symptom research is affected from modality-specific measurement variability (Moriarity and Alloy, 2021) and in our study this is aggravated through the added variability unique to the method of symptom assessment (Möller, 2000). Future studies would benefit from inclusion of studies with the same questionnaire and disaggregated symptom measures.

Second, the combination of clinical and general population samples poses unique challenges. The application of a clinical depression measure in the UK Biobank study could have resulted in potential floor effects for some symptoms while specific selection of MDD patients into MARS and STAR*D studies could have resulted in ceiling effects for core symptoms of depressed mood and anhedonia as these are required for a diagnosis. Selection of clinical populations in network studies can also result in Berkson/collider bias (de Ron et al., 2019), which can induce negative correlations. This again warrants replication of our results in independent samples.

Third, PRSs are based on GWAS with highly diverging samples sizes as a large number of individuals were included in the GWAS for BMI and CRP (>200 thousand individuals) and smaller numbers of individuals (∼8 thousand individuals) for IL-6, IL-10, and TNF-α.

Consistency of effect sizes following the PRS-CS approach was also larger for CRP and BMI as compared to IL-6, IL-10, and TNF-α. This is likely to have shifted the balance of statistical power towards detection of PRS-symptom associations to BMI and CRP rather than IL-6, IL-10, and TNF-α. Therefore, our findings require replication once larger individual cytokine GWAS become available.

## Conclusion

The present investigation studied associations between four major pro- and anti-inflammatory markers, BMI, and depressive symptoms by applying network analysis across one large general population and two patient samples. Defining immuno-metabolic markers using polygenic risk scores expanded previous reports by suggesting direct associations of genetic/lifetime predisposition to immune-metabolic markers with depressive symptoms and provided evidence for temporality of association. Despite methodological restrictions of the presented approach, we observed associations of polygenic risk for CRP with changes in appetite and fatigue, for TNF-α with fatigue, and similar associations for BMI. These findings align with recent conceptualisations of an immuno-metabolic subgroup of depressed patients characterised by atypical, neurovegetative symptom profiles. Results can inform future clinical trials of anti-inflammatory approaches by prioritising these patients for selection into clinical trials.

## Supporting information

STROBE checklist

## Data Availability

Data from original studies is not openly available, but can be requested; see details in Supplementary Table 7. GWAS summary data for IL-6, IL-10, and TNF-α is openly available from the original publication by Ahola-Olli and colleagues (2017), for BMI from the GIANT consortium, and can be requested for CRP from the CHARGE inflammation working group. We provide analysis scripts and estimated network matrices (including bootstrapped network matrices) on the Open Science Platform (OSF) under https://osf.io/q4vw9/.

## DECLARATION OF INTERESTS

The authors do not have any competing interests.

## FUNDING

This study is funded by the Max Planck Institute of Psychiatry. NK, NR, and SM are supported by the International Max Planck Research School of Translational Psychiatry (IMPRS-TP). NR received funding from the Bavarian Ministry of Economic Affairs, Regional Development and Energy (BayMED, PBN_MED-1711-0003). GMK acknowledges funding support from the Wellcome Trust (grant code: 201486/Z/16/Z), the MQ: Transforming Mental Health (grant code: MQDS17/40), the Medical Research Council, UK (grant code: MC_PC_17213 and grant code: MR/S037675/1), and the BMA Foundation (J Moulton grant 2019). JA received support by a NARSAD Young Investigator Grant from Brain and Behavior Research Foundation.

## ACKNOWLEDGMENTS

We are grateful to all original authors, technical assistants and patients who contributed to the MARS study. We are grateful for the National Institute of Mental Health (NIMH) and the NIMH Repository and Genomics Resource (NRGR) for the possibility of analysing the STAR*D data. We are also grateful to the original STAR*D authors, and particularly for the contributions of all patients and families who participated in the study. Data were obtained from the limited access datasets distributed from the NIH-supported ‘Sequenced Treatment Alternatives to Relieve Depression’ (STAR*D). The study was supported by NIMH Contract No. N01MH90003 to the University of Texas Southwestern Medical Center. The ClinicalTrials.gov identifier is NCT00021528. This research has been conducted using the UK Biobank Resource. We are grateful for all scientists and participants who made this large-scale effort and resource possible.

## Notes

### Competing Interest Statement

The authors have declared no competing interest.

### Author Declarations

We report secondary analyses for data from existing studies. These studies have received ethics committee approval, which we have described as follows in the manuscript: "MARS received local ethics approval from Ludwig Maximilians University Munich (Hennings et al., 2009). STAR*D received ethics approval from 14 participating institutional review boards, a National Coordinating Center, a Data Coordinating Center, and the Data Safety and Monitoring Board at the National Institute of Mental Health (Rush et al., 2006, 2004). The UK Biobank study received ethics approval from North West Centre for Research Ethics Committee and Human Tissue Authority research tissue bank (Bycroft et al., 2018); this project was approved under project no. 26999. All three studies collected informed consent from participants prior to study participation."

### Summary of Updates

Revised manuscript, abstract, supplementary material, and figure 1.

