## Supplementary Material for "Polygenic risk for immuno-metabolic markers and specific depressive symptoms: A multi-sample network analysis study"

### SUPPLEMENTARY METHODS

#### Genotyping, quality control and imputation

##### MARS

Genotyping in the MARS study was conducted using three genotyping arrays, Illumina 610k, Illumina Omni Express and Illumina GSA. The Illumina 610k array was used for individuals recruited between 1996 and 2015 (Interquartile range [IQR]: 2001-2006), the Illumina Omni Express between 1996 and 2013 (IQR: 2008-2010), and the Illumina GSA array between 2011 and 2015 (IQR: 2012-2015); see Supplementary Figure 1. Quality control was performed for each genotyping batch individually in PLINK (Purcell et al., 2007). We removed SNPs with a callrate below 98%, a minor allele frequency below 1% or deviation from Hardy-Weinberg-Equilibrium (p-value < 1e^-5^). Furthermore, individuals presenting with callrates below 98% were excluded. Samples with outlying multi-dimensional scaling components (more than four standard deviations below/above the mean on any of the first 10 axes) as well as heterozygosity outliers (more than four standard deviations below/above the mean heterozygosity) were removed. We only included unrelated individuals in the analysis. We used shapeit2 (Delaneau et al., 2012) for haplotype phasing and impute2 (Howie et al., 2009) with the 1,000 Genomes Phase III reference samples. After imputation, only SNPs with an info-metric above 0.6, a minor allele frequency above 1% and no deviation from Hardy-Weinberg-Equilibrium (p>e^-5^) were retained in the analysis. Genotype probabilities were recoded into best-guessed genotypes using a hard-call threshold of 0.1, i.e. individual genotypes presenting with probabilities below 0.9 were set as missing. The final datasets contained 548 samples with SNPs 9,596,975 SNPs (Illumina 610k), 284 samples with 6,765,193 SNPs (Illumina Omni Express) and 226 samples with 9,572,313 SNPs (Illumina GSA).

##### Supplementary Figure 1. Distribution of time of recruitment into MARS per genotyping array

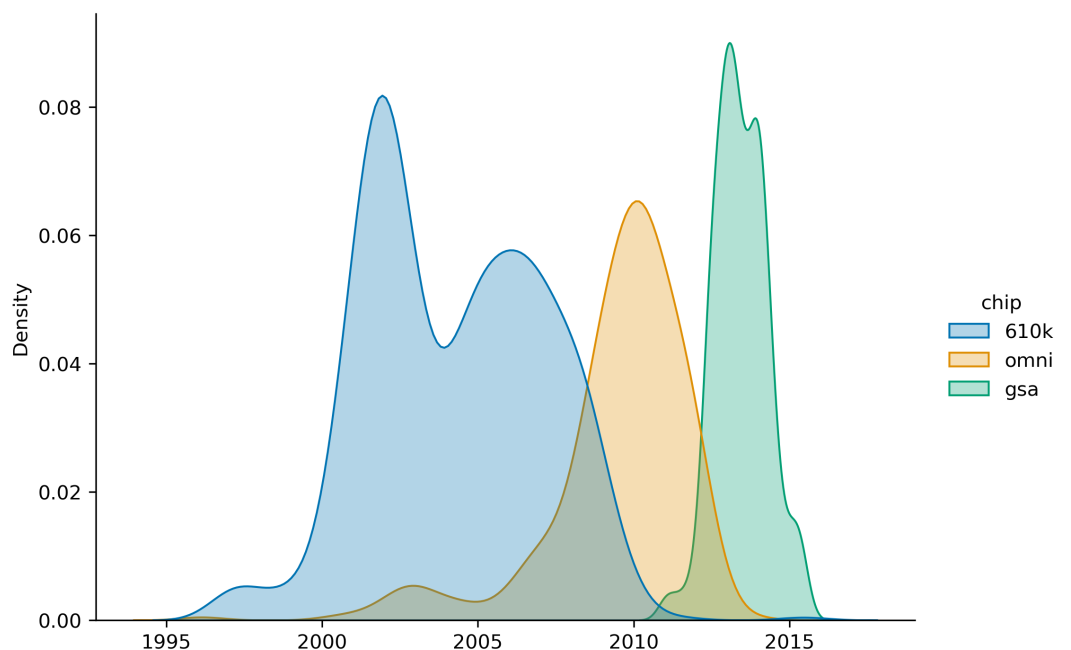

*Note*: The x-axis of this density plot displays time of recruitment into the MARS study with differential colour coding for Illumina 610k (blue), Illumina OmniExpress (orange), and Illumina GSA (green) genotyping arrays.

##### STAR*D

Genotyping was conducted using the Affymetrix Human Mapping 500K Array Set (n=979) and the Affymetrix Genome-Wide Human SNP Array 5.0 (n=969), which displayed a concordance of >99% as assessed by twelve participants genotyped using both platforms. Further details are described by Garriock *and colleagues* (2010).

Only unrelated individuals of white ethnicity were included. Quality control was performed in PLINK (Purcell et al., 2007). We removed SNPs with a callrate below 98%, a minor allele frequency below 1% or deviation from Hardy-Weinberg-Equilibrium (p-value < 1e^-5^). Furthermore, individuals presenting with callrates below 98% were excluded. Samples with outlying multi-dimensional scaling components (more than four standard deviations below/above the mean on any of the first 10 axes) as well as heterozygosity outliers (more than four standard deviations below/above the mean heterozygosity) were removed. We used shapeit2 (Delaneau et al., 2012) for haplotype phasing and impute2 (Howie et al., 2009) with the 1,000 Genomes Phase III reference samples. After imputation, only SNPs with an info-metric above 0.6, a minor allele frequency above 1% and no deviation from Hardy-Weinberg-Equilibrium (p>e^-5^) were retained in the analysis. Genotype probabilities were recoded into best-guessed genotypes using a hard-call threshold of 0.1, i.e. individual genotypes presenting with probabilities below 0.9 were set as missing. The final dataset included 1,143 individuals and 8,267,857 SNPs.

##### UK Biobank

The imputed UK Biobank genetic data (version 3) for ca 488,000 participants was used as genotyped on the UK BiLEVE Axiom Array (~45,000 samples) or the Affymetrix UK Biobank Axiom Array. Quality Control before imputation and the imputation were performed centrally by the Welcome Trust Centre for Human Genetics as described by Bycroft *and colleagues* (2018). In the post-imputation quality control, we excluded individuals with non-British ancestry and individuals related up to the third degree resulting in a final dataset of 342,081 individuals. Per-variant quality control was performed using QCTOOL (<https://enkre.net/cgi-bin/code/qctool/dir?ci=trunk>) to exclude SNPs minor allele frequency < 1% , imputation score < 60%, genotyping missingness > 2%, and Hardy-Weinberg Equilibrium test p-value < 0.000001.

#### Depressive symptom assessment

##### Supplementary Table 1: Symptom availability and selection across samples

| **HAM-D (MARS & STAR*D)** | | | **PHQ-9 (UK Biobank)** | | | | |
| --- | --- | --- | --- | --- | --- | --- | --- |
| **Item** | **Rating description** | **Label in this study** | **Item** | **Question** | **Rating** | **Label in this study** | **UKB field code** |
| 1* | Depressed Mood (Sad, blue, gloomy, down) | Depressed mood | 1* | Over the last 2 weeks, how often have you been bothered by any of the following problems? | Little interest or pleasure in doing things | Anhedonia | 20514 |
| 2 | Guilt Feelings and Delusions |  | 2* | Over the last 2 weeks, how often have you been bothered by any of the following problems? | Feeling down, depressed, or hopeless | Depressed mood | 20510 |
| 3* | Suicide | Suicidality | 3* | Over the last 2 weeks, how often have you been bothered by any of the following problems? | Trouble falling or staying asleep, or sleeping too much | Sleep problems | 20517 |
| 4* | Initial Insomnia | Sleep problems | 4* | Over the last 2 weeks, how often have you been bothered by any of the following problems? | Feeling tired or having little energy | Fatigue | 20519 |
| 5* | Middle Insomnia (between 12:00 and 4:00a.m.) | Sleep problems | 5* | Over the last 2 weeks, how often have you been bothered by any of the following problems? | Poor appetite or overeating | Changes in appetite | 20511 |
| 6* | Delayed Insomnia (after 4:00a.m.) | Sleep problems | 6 | Over the last 2 weeks, how often have you been bothered by any of the following problems? | Feeling bad about yourself or that you are a failure or have let yourself or your family down |  | 20507 |
| 7* | Work and Interests (Apathy: loss of interest in work,  hobbies, social life. Anhedonia: unable to feel  pleasure) | Anhedonia | 7 | Over the last 2 weeks, how often have you been bothered by any of the following problems? | Trouble concentrating on things, such as reading the newspaper or watching television |  | 20508 |
| 8* | Retardation (Psychomotor slowing of thought,  speech, and movement) | Psychomotor changes | 8* | Over the last 2 weeks, how often have you been bothered by any of the following problems? | Moving or speaking so slowly that other people could have noticed? Or the opposite - being so fidgety or restless that you have been moving around a lot more than usual | Psychomotor changes | 20518 |
| 9* | Agitation (May co-exist with retardation) | Psychomotor changes | 9* | Over the last 2 weeks, how often have you been bothered by any of the following problems? | Thoughts that you would be better off dead or of hurting yourself in some way | Suicidality | 20513 |
| 10 | Psychic Anxiety (Includes feeling tense, irritable,  apprehensive, fearful, phobic, or panic attacks) |  |  |  |  |  |  |
| 11 | Somatic Anxiety (Physiological concomitants of  anxiety such as: fainting, tinnitus, blurred vision,  headache, tremor, sweating, flushing, hyperventilation,  palpitations, indigestion, belching, diarrhea, increased  urinary frequency) |  |  |  |  |  |  |
| 12* | Appetite | Changes in appetite |  |  |  |  |  |
| 13* | Somatic Energy | Fatigue |  |  |  |  |  |
| 14 | Libido |  |  |  |  |  |  |
| 15 | Hypochondriasis (Preoccupation with physical  health) (Do not rate concern about physical fitness  unless outside the norm) |  |  |  |  |  |  |
| 16 | Weight Loss (Within the Last Week) |  |  |  |  |  |  |
| 17 | Loss of Insight |  |  |  |  |  |  |

*Note*: *These items were selected for joint analyses in the present study. UKB=UK Biobank.

##### Supplementary Table 2. Symptom coding

|  | **HAM-D (MARS & STAR*D** | | **PHQ-9 (UK Biobank)** | |
| --- | --- | --- | --- | --- |
| **Symptom** | **Items** | **Coding** | **Items** | **Coding** |
| Depressed mood | 1 | - | 2 | - |
| Anhedonia | 7 | - | 1 | - |
| Sleep problems | 4, 5, 6 | Sum of 4, 5, & 6 | 3 | - |
| Fatigue | 13 | - | 4 | - |
| Changes in appetite | 12 | - | 5 | - |
| Psychomotor changes | 8, 9 | Max. of 8 & 9 | 8 | - |
| Suicidality | 3 | - | 9 | - |

*Note*: Symptoms with “-“ indicated in coding columns have retained original coding from HAM-D and PHQ-9 questionnaires. Max=Maximum.

#### Polygenic risk score computation

PRS-CS requires specification of the global shrinkage parameter *ϕ*, which can be pre-specified, selected using grid search, or learnt from GWAS summary data (Ge et al., 2019). We pre-specified *ϕ* because grid search selection of tuning parameters should ideally be done in independent data, which was not available, and automatic learning from GWAS summary data (referred to as PRS-CS AUTO in the literature) has shown sub-optimal performance for PRSs of psychiatric traits (Ni et al., 2020). Pre-specification was based on suggested values for highly polygenic (*ϕ*=1e^-2^) and less polygenic (*ϕ*=1e^-4^) traits (see <https://github.com/getian107/PRScs>). We set *ϕ*=1e^-4^ for CRP, IL-6, IL-10, and TNF-α as these are relatively less polygenic compared to BMI, for which we set *ϕ*=1e^-2^, and to assure PRSs are based on more unique genetic contributions for each inflammatory marker as signalling pathways are interrelated (Rose-John, 2018; Steensberg et al., 2003). The impact of pre-specification on resulting PRSs was also evaluated (see next section & Supplementary Table 6).

#### Polygenic risk score evaluation

##### Supplementary Table 3. Number of SNPs following PRS-CS shrinkage

|  |  | **MARS** |  |  |  |
| --- | --- | --- | --- | --- | --- |
| **PRS** | **610k** | **GSA** | **Omni** | **STAR*D** | **UK Biobank** |
| CRP | 1,104,080 | 1,096,869 | 986,135 | 1,002,153 | 1,095,433 |
| IL-6 | 1,098,387 | 1,090,507 | 980,444 | 996,158 | 1,089,691 |
| IL10 | 1,098,316 | 1,090,471 | 980,389 | 996,129 | 1,089,630 |
| TNF-α | 1,096,208 | 1,088,458 | 979,283 | 994,709 | 1,087,617 |
| BMI | 1,019,852 | 1,011,770 | 911,738 | 926,135 | 1,012,569 |

##### Supplementary Table 4. Proportion of overlapping SNPs between samples following PRS-CS shrinkage

|  |  | **MARS** |  |  |  |
| --- | --- | --- | --- | --- | --- |
| **PRS** | **610k** | **GSA** | **Omni** | **STAR*D** | **UK Biobank** |
| ***CRP*** |  |  |  |  |  |
| MARS: 550k | 1 | 0.99 | 0.892 | 0.906 | 0.991 |
| MARS: GSA | 0.996 | 1 | 0.893 | 0.907 | 0.989 |
| MARS: Omni | 0.999 | 0.994 | 1 | 0.922 | 0.992 |
| STAR*D | 0.998 | 0.993 | 0.907 | 1 | 0.991 |
| UK Biobank | 0.999 | 0.99 | 0.893 | 0.907 | 1 |
| ***IL-6*** |  |  |  |  |  |
| MARS: 550k | 1 | 0.99 | 0.892 | 0.905 | 0.991 |
| MARS: GSA | 0.997 | 1 | 0.893 | 0.907 | 0.989 |
| MARS: Omni | 0.999 | 0.994 | 1 | 0.922 | 0.992 |
| STAR*D | 0.998 | 0.993 | 0.907 | 1 | 0.991 |
| UK Biobank | 0.999 | 0.99 | 0.893 | 0.906 | 1 |
| ***IL-10*** |  |  |  |  |  |
| MARS: 550k | 1 | 0.99 | 0.892 | 0.905 | 0.991 |
| MARS: GSA | 0.997 | 1 | 0.893 | 0.907 | 0.989 |
| MARS: Omni | 0.999 | 0.994 | 1 | 0.922 | 0.992 |
| STAR*D | 0.998 | 0.993 | 0.907 | 1 | 0.991 |
| UK Biobank | 0.999 | 0.99 | 0.893 | 0.906 | 1 |
| ***TNF-α*** |  |  |  |  |  |
| MARS: 550k | 1 | 0.99 | 0.892 | 0.906 | 0.991 |
| MARS: GSA | 0.997 | 1 | 0.894 | 0.907 | 0.989 |
| MARS: Omni | 0.999 | 0.994 | 1 | 0.922 | 0.992 |
| STAR*D | 0.998 | 0.993 | 0.908 | 1 | 0.991 |
| UK Biobank | 0.999 | 0.99 | 0.893 | 0.907 | 1 |
| ***BMI*** |  |  |  |  |  |
| MARS: 550k | 1 | 0.989 | 0.893 | 0.907 | 0.991 |
| MARS: GSA | 0.997 | 1 | 0.895 | 0.909 | 0.99 |
| MARS: Omni | 0.999 | 0.994 | 1 | 0.923 | 0.992 |
| STAR*D | 0.998 | 0.993 | 0.908 | 1 | 0.992 |
| UK Biobank | 0.999 | 0.989 | 0.894 | 0.907 | 1 |

*Note*: Proportions differ above versus below the diagonal. Proportions above the diagonal reflect the proportion of available SNPs of the sample noted in the column that was also present in the sample noted in the row while proportions below the proportion of available SNPs of the sample noted in the row that was also present in the sample noted in the column.

##### Supplementary Table 5. Effect size correlations of common SNPs between samples following PRS-CS shrinkage

|  |  | **MARS** |  |  |  |
| --- | --- | --- | --- | --- | --- |
| **PRS** | **610k** | **GSA** | **Omni** | **STAR*D** | **UK Biobank** |
| ***CRP*** |  |  |  |  |  |
| MARS: 550k | 1 |  |  |  |  |
| MARS: GSA | 0.76 | 1 |  |  |  |
| MARS: Omni | 0.71 | 0.7 | 1 |  |  |
| STAR*D | 0.73 | 0.72 | 0.69 | 1 |  |
| UK Biobank | 0.75 | 0.75 | 0.69 | 0.73 | 1 |
| ***IL-6*** |  |  |  |  |  |
| MARS: 550k | 1 |  |  |  |  |
| MARS: GSA | 0.42 | 1 |  |  |  |
| MARS: Omni | 0.41 | 0.42 | 1 |  |  |
| STAR*D | 0.41 | 0.42 | 0.41 | 1 |  |
| UK Biobank | 0.43 | 0.42 | 0.42 | 0.41 | 1 |
| ***IL-10*** |  |  |  |  |  |
| MARS: 550k | 1 |  |  |  |  |
| MARS: GSA | 0.45 | 1 |  |  |  |
| MARS: Omni | 0.43 | 0.43 | 1 |  |  |
| STAR*D | 0.43 | 0.44 | 0.42 | 1 |  |
| UK Biobank | 0.45 | 0.46 | 0.43 | 0.43 | 1 |
| ***TNF-α*** |  |  |  |  |  |
| MARS: 550k | 1 |  |  |  |  |
| MARS: GSA | 0.42 | 1 |  |  |  |
| MARS: Omni | 0.41 | 0.42 | 1 |  |  |
| STAR*D | 0.42 | 0.42 | 0.41 | 1 |  |
| UK Biobank | 0.43 | 0.43 | 0.41 | 0.44 | 1 |
| ***BMI*** |  |  |  |  |  |
| MARS: 550k | 1 |  |  |  |  |
| MARS: GSA | 0.8 | 1 |  |  |  |
| MARS: Omni | 0.79 | 0.79 | 1 |  |  |
| STAR*D | 0.79 | 0.79 | 0.79 | 1 |  |
| UK Biobank | 0.8 | 0.8 | 0.79 | 0.79 | 1 |

*Note*: Pearson’s correlation coefficients were used.

##### Supplementary Table 6. PRS intercorrelations using different PRS-CS settings for the hyperparameter ϕ

|  |  | **MARS** |  |  |  |
| --- | --- | --- | --- | --- | --- |
| **PRS** | **610k** | **GSA** | **Omni** | **STAR*D** | **UK Biobank** |
| ***CRP (ϕ=1e^-4^)*** |  |  |  |  |  |
| *ϕ*=1e^-6^ | 0.86 | 0.87 | 0.84 | 0.87 | 0.87 |
| *ϕ* with PRS-CS auto | 0.98 | 0.98 | 0.98 | 0.97 | 0.98 |
| *ϕ*=1 | 0.63 | 0.7 | 0.61 | 0.59 | 0.63 |
| ***IL-6 (ϕ=1e^-4^)*** |  |  |  |  |  |
| *ϕ*=1e^-6^ | 0.69 | 0.73 | 0.71 | 0.69 | 0.7 |
| *ϕ* with PRS-CS auto | 0.92 | 0.92 | 0.91 | 0.92 | 0.92 |
| *ϕ*=1 | 0.69 | 0.77 | 0.75 | 0.71 | 0.71 |
| ***IL-10 (ϕ=1e^-4^)*** |  |  |  |  |  |
| *ϕ*=1e^-6^ | 0.59 | 0.57 | 0.66 | 0.66 | 0.61 |
| *ϕ* with PRS-CS auto | 0.93 | 0.93 | 0.93 | 0.91 | 0.92 |
| *ϕ*=1 | 0.73 | 0.72 | 0.76 | 0.7 | 0.69 |
| ***TNF-α (ϕ=1e^-4^)*** |  |  |  |  |  |
| *ϕ*=1e^-6^ | 0.69 | 0.73 | 0.66 | 0.67 | 0.69 |
| *ϕ* with PRS-CS auto | 0.91 | 0.93 | 0.91 | 0.92 | 0.92 |
| *ϕ*=1 | 0.72 | 0.7 | 0.74 | 0.7 | 0.71 |
| ***BMI (ϕ=1e^-2^)*** |  |  |  |  |  |
| *ϕ*=1e^-6^ | 0.54 | 0.56 | 0.51 | 0.47 | 0.48 |
| *ϕ* with PRS-CS auto | 0.82 | 0.86 | 0.85 | 0.83 | 0.84 |
| *ϕ*=1 | 0.91 | 0.93 | 0.93 | 0.91 | 0.92 |

##### Sensitivity analysis in MARS genotyping array subgroups

To evaluate potential residual effects of the different genotyping arrays used in MARS, we performed further sensitivity analysis.

First, Supplementary Figure 2 demonstrates highly similar PRS distributions across PRS phenotypes. Second, we estimated an additional network of depressive symptoms and PRSs, as done in main analyses, but additionally including the genotyping array as a node in the network. As the genotyping array is a categorical variable, which can currently not be estimated using FGL or unregularized model search estimation, we used a mixed graphical model for this sensitivity analysis. Results of this model are displayed in Supplementary Figure 3 and do not show any remaining edges between the genotyping array and PRSs suggesting that the PRS-symptom edges should not be biased by the genotyping array used. However, we did observe significant edges between the genotyping chip node and several symptoms in the range of 0 to 0.19. As genotyping arrays differed across a relatively long study period in MARS (cf. Supplementary Figure 1), this suggests potential differences in the assessment of the Hamilton Depression Rating Scale, an observer-rated scale for depression, over time. Nonetheless, overlapping PRS distributions and absence of PRS-genotyping array edges justifies combination of MARS subsamples for further analyses.

##### Supplementary Figure 2. PRS distribution by genotyping chip in MARS

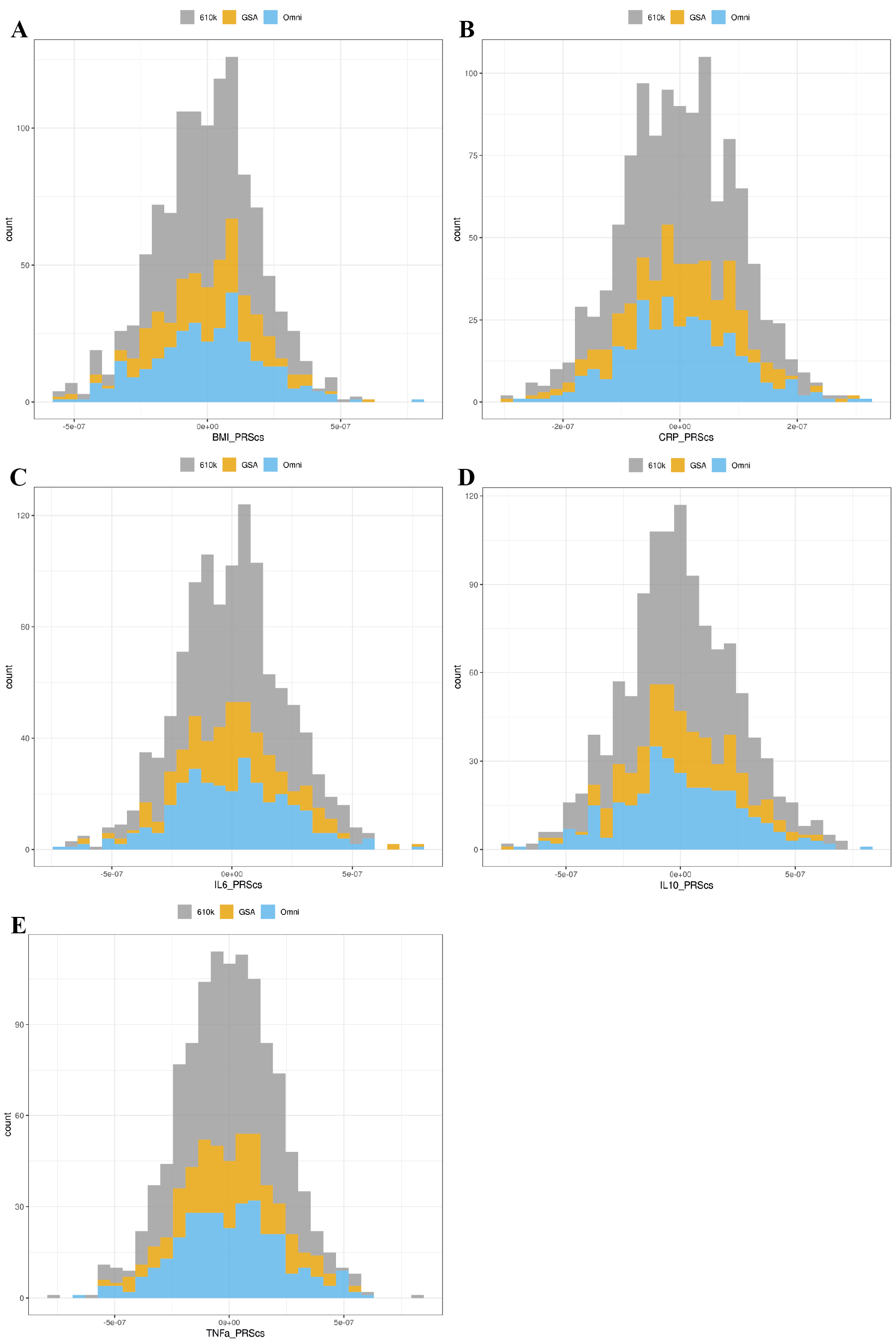

*Note*: PRS distributions are shown for BMI (A), CRP (B), IL-6 (C), IL-10 (D), TNF-α (E) with grey indicating the Illumina 610k, orange indicating the Illumina GSA, and blue indicating the Illumina OmniExpress genotyping array.

##### Supplementary Figure 3. Mixed graphical model between symptoms, PRSs and genotyping array in MARS

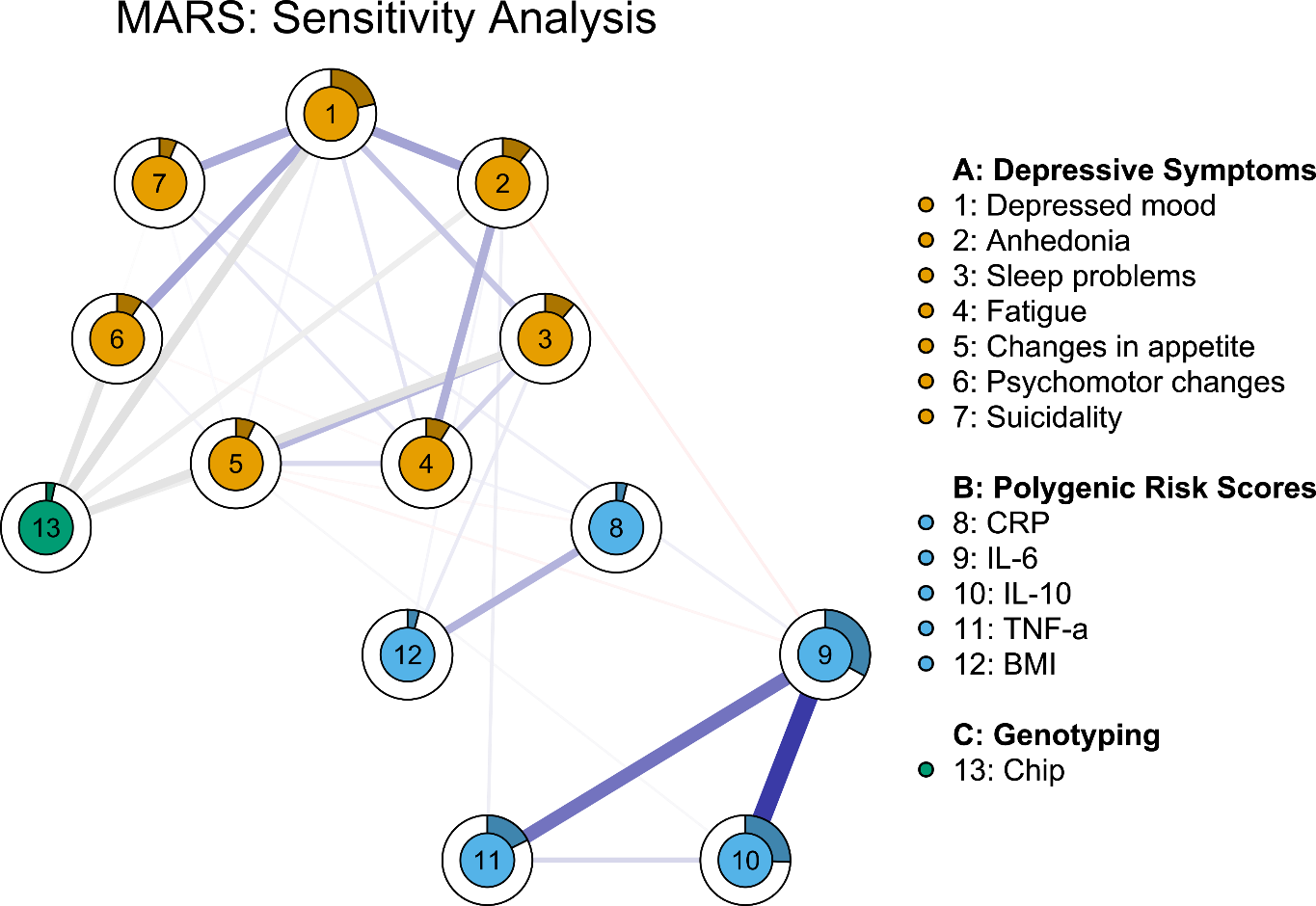

*Note*: Networks are visualised with the *qgraph* package. Blue lines indicate positive and red lines negative associations, respectively, with larger associations displayed with thicker lines. Circles around nodes display node predictability, which can be interpreted similar to explained variance. Maximum size of edge associations is 0.55. Changes in appetite and sleep problems are measured as composite symptoms in UK Biobank, but as loss of appetite and insomnia in MARS and STAR*D samples.

#### Availability of data and materials

##### Supplementary Table 7. Availability of original data

| Cohort | Means to obtain original data |
| --- | --- |
| MARS | Request data by contacting:  Dr Tanja Brückl  Max Planck Institute of Psychiatry, Munich, Germany  |
| STAR*D | Formal request through the National Institute of Mental Health (NIMH) and the NIMH Repository and Genomics Resource (NRGR) via <https://www.nimhgenetics.org/request-access/how-to-request-access> |
| UK Biobank | Formal request through the UK Biobank website via <https://www.ukbiobank.ac.uk/enable-your-research/apply-for-access> |

### SUPPLEMENTARY RESULTS

#### Fused Graphical LASSO (FGL) estimation

##### Supplementary Figure 4. Bootstrapped 95% quantile intervals of symptom-symptom edges using FGL estimation

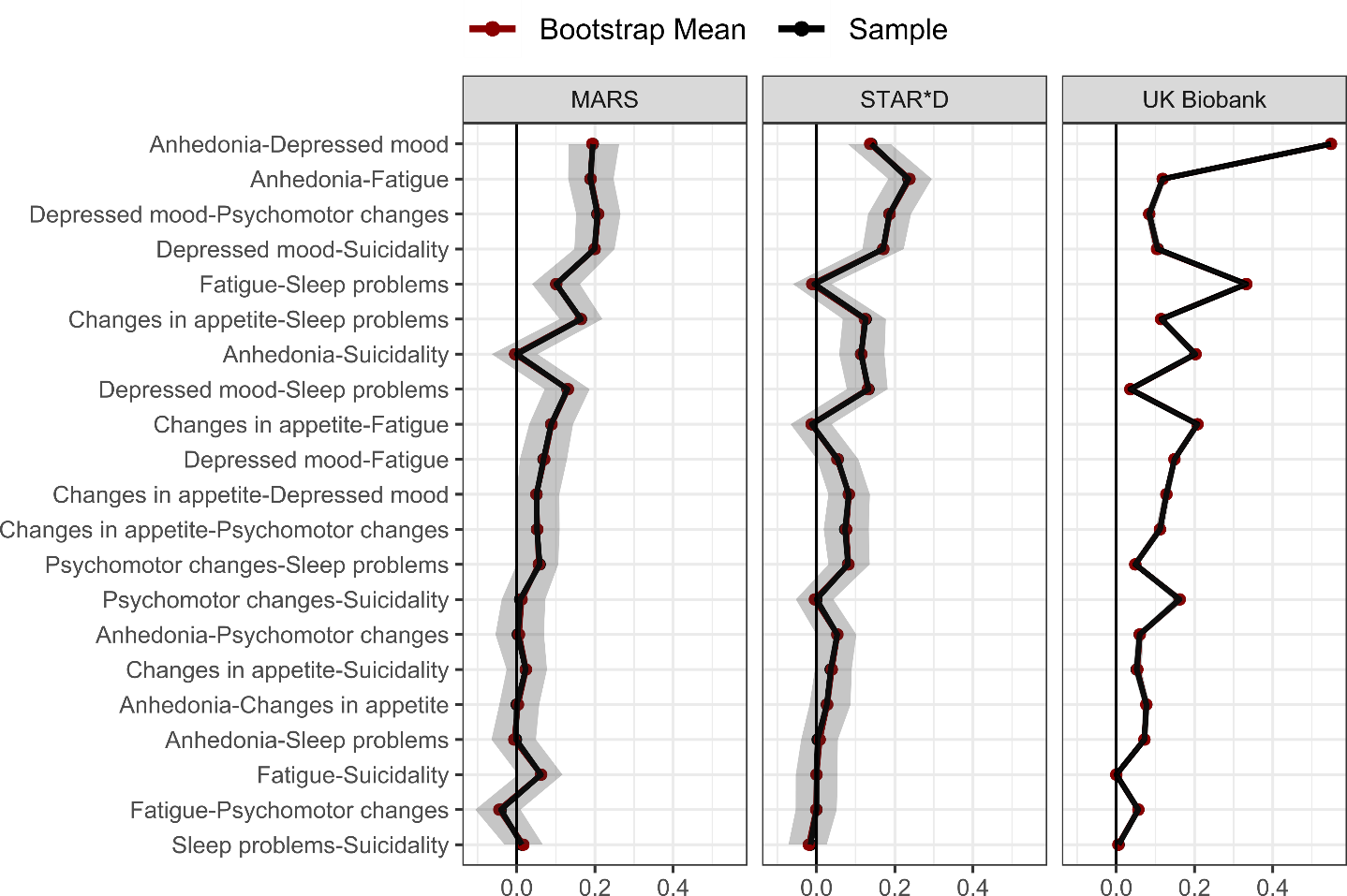

*Note*: Bootstrapped 95% quantile intervals (i.e., 95% of the distribution of raw bootstrapped edge estimates) are highlighted as shaded area for each edge. Black points indicate the raw FGL sample estimate while red points indicate the raw bootstrapped mean estimate. Edges are indicated on the y-axis and sorted by mean edge size across samples in descending order.

##### Supplementary Figure 5. Bootstrapped 95% quantile intervals of PRS-PRS edges using FGL estimation

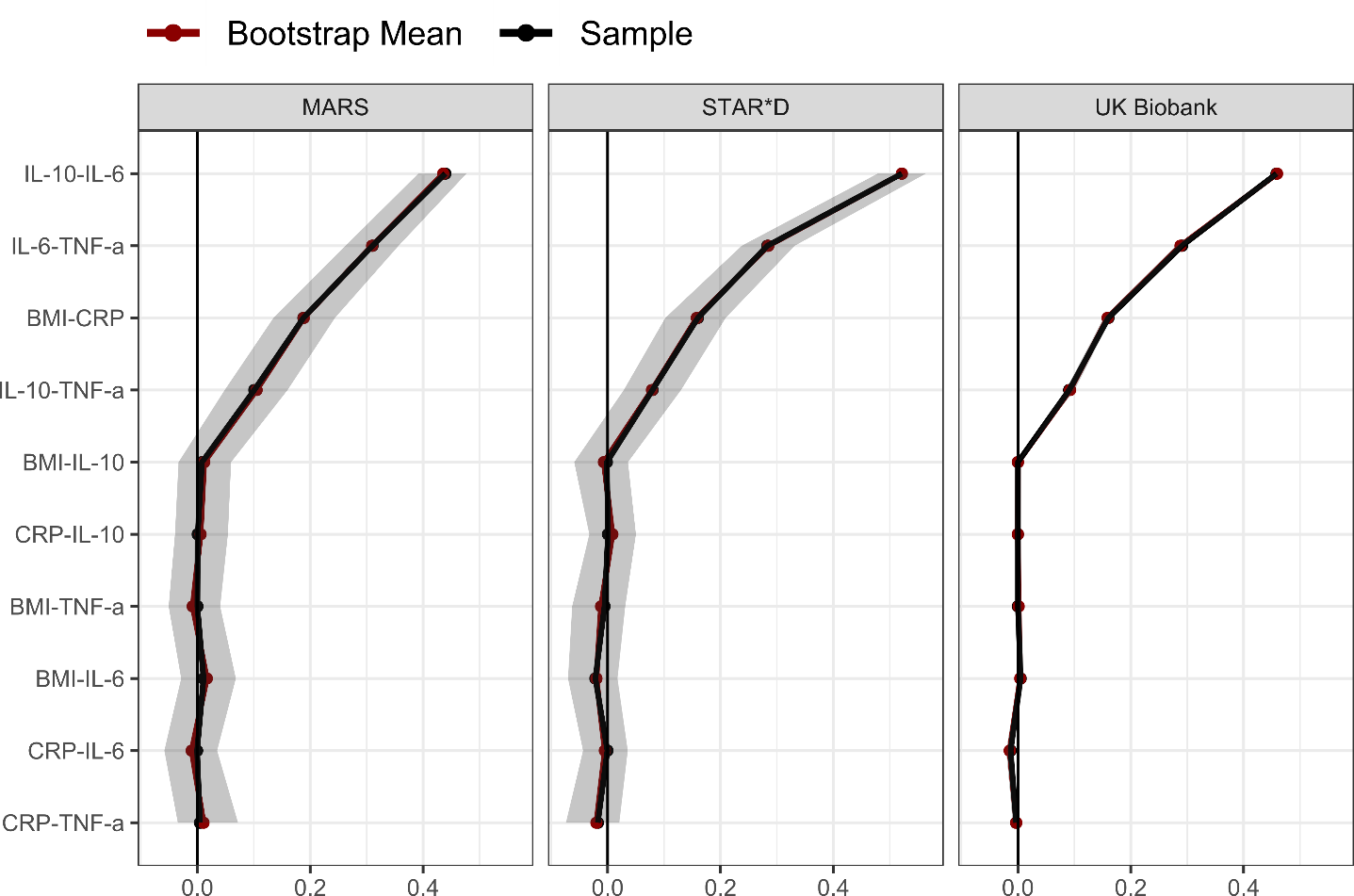

*Note*: Bootstrapped 95% quantile intervals (i.e., 95% of the distribution of raw bootstrapped edge estimates) are highlighted as shaded area for each edge. Black points indicate the raw FGL sample estimate while red points indicate the raw bootstrapped mean estimate. Edges are indicated on the y-axis and sorted by mean edge size across samples in descending order.

#### Unregularized model search estimation

##### Supplementary Figure 6. Estimated unregularized model search networks across samples

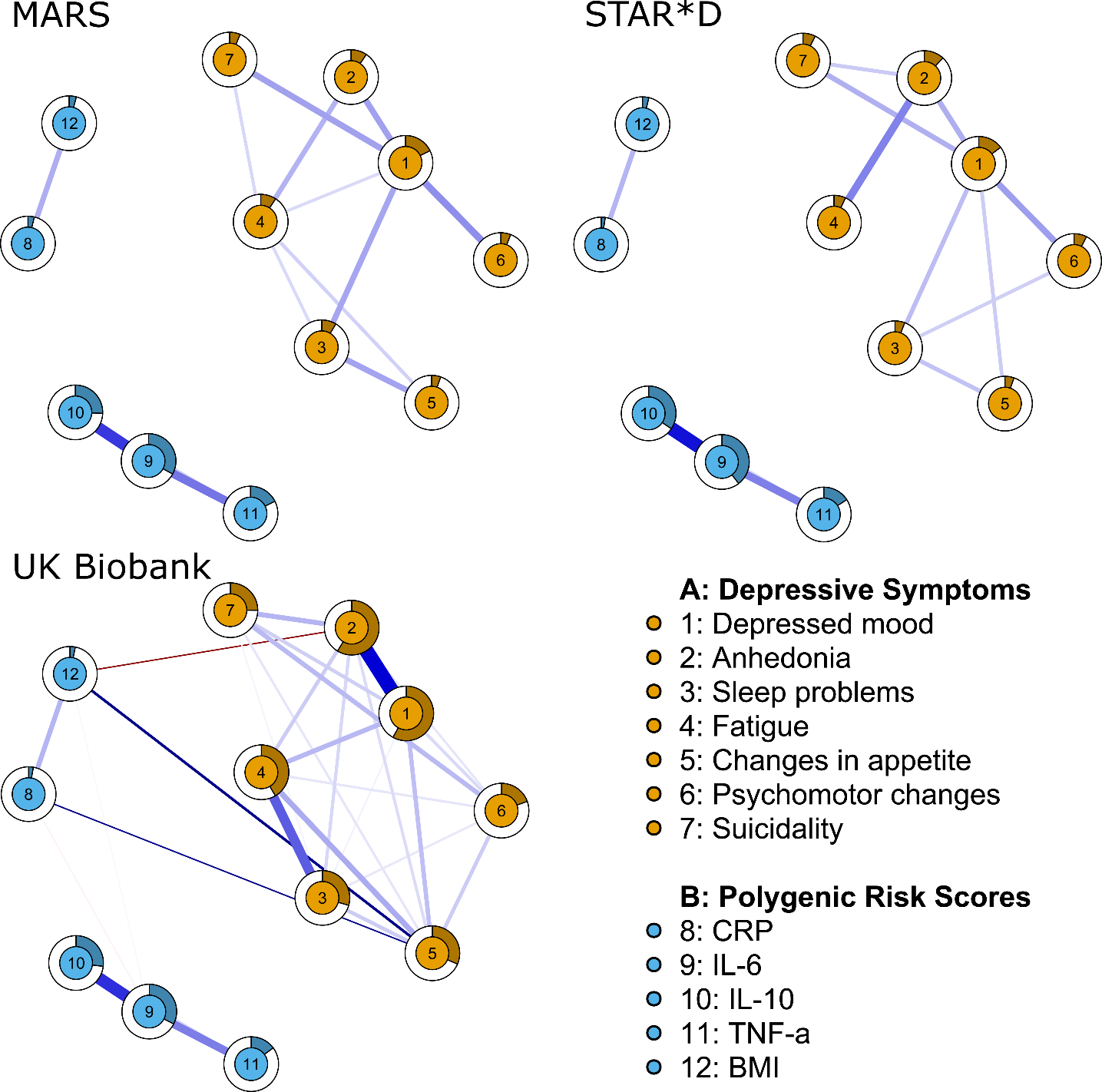

*Note*: Networks are visualised with the *qgraph* package. Blue lines indicate positive and red lines negative associations, respectively, with larger associations displayed with thicker lines. Circles around nodes display node predictability, which can be interpreted similar to explained variance. Maximum size of edge associations is 0.55. As the primary focus of this investigation was to identify consistent PRS-symptom associations, we manually unfaded edges between PRSs and symptoms if these edges met quality criterion 3 (see Table 2). Changes in appetite and sleep problems are measured as composite symptoms in UK Biobank, but as loss of appetite and insomnia in MARS and STAR*D samples.

##### Supplementary Figure 7. Bootstrapped 95% quantile intervals of symptom-symptom edges using unregularized model search estimation

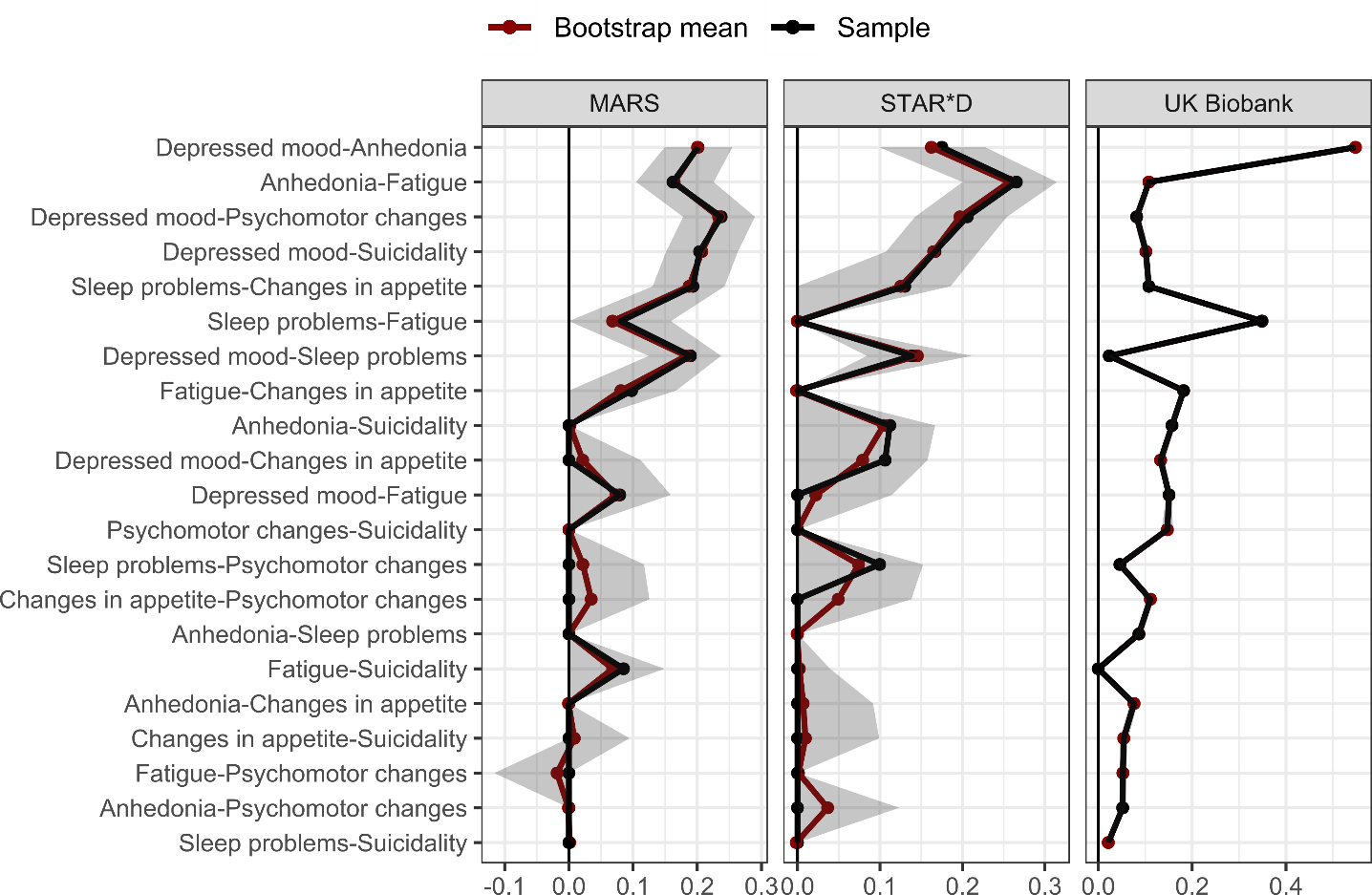

*Note*: Bootstrapped 95% quantile intervals (i.e., 95% of the distribution of raw bootstrapped edge estimates) are highlighted as shaded area for each edge. Black points indicate the raw FGL sample estimate while red points indicate the raw bootstrapped mean estimate. Edges are indicated on the y-axis and sorted by mean edge size across samples in descending order.

##### Supplementary Figure 8. Bootstrapped 95% quantile intervals of PRS-symptom edges using unregularised model search estimation

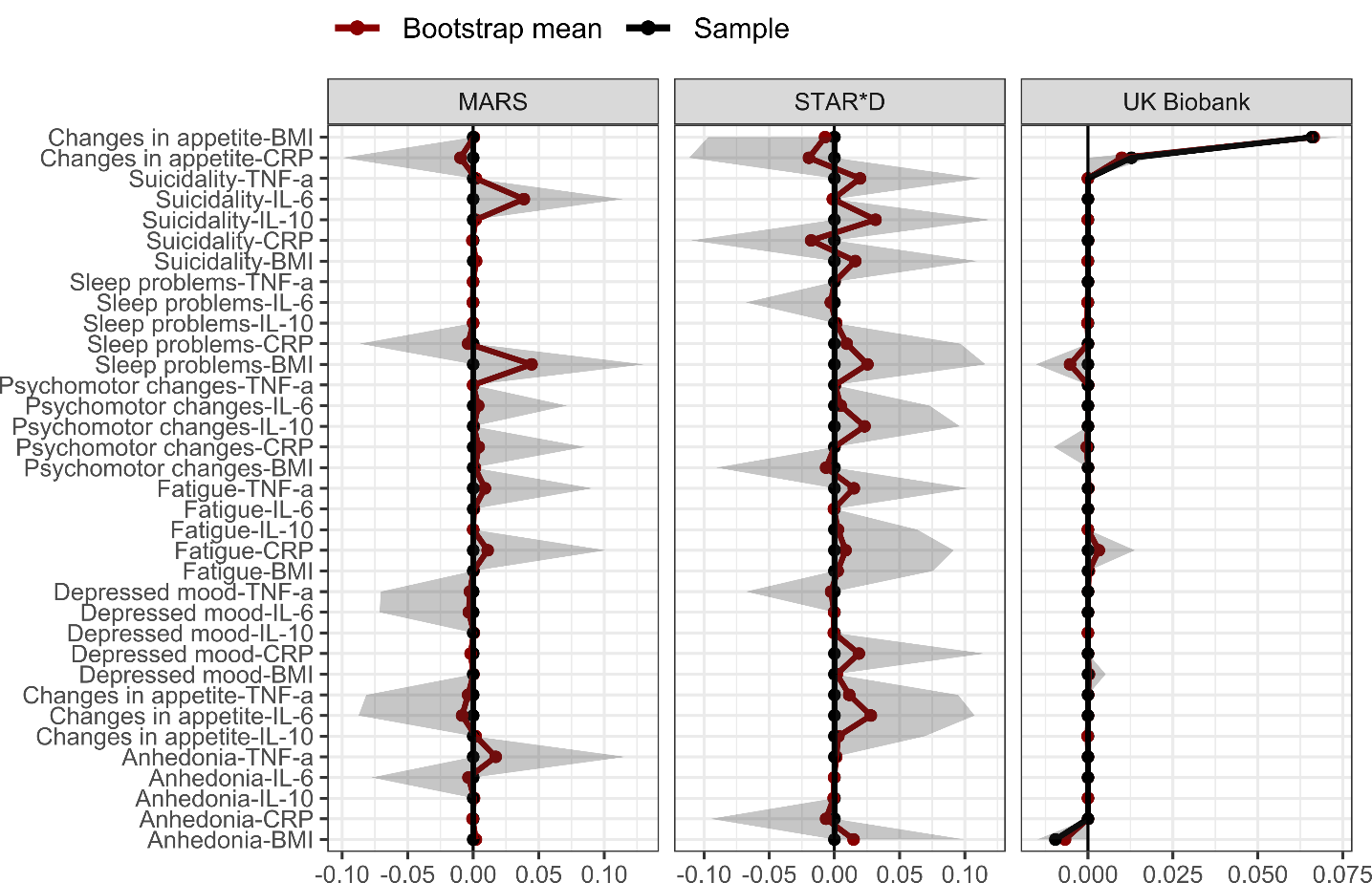

*Note*: Bootstrapped 95% quantile intervals (i.e., 95% of the distribution of raw bootstrapped edge estimates) are highlighted as shaded area for each edge. Black points indicate the raw FGL sample estimate while red points indicate the raw bootstrapped mean estimate. Edges are indicated on the y-axis and sorted by mean edge size across samples in descending order.

##### Supplementary Figure 9. Bootstrapped 95% quantile intervals of PRS-PRS edges using unregularised model search estimation

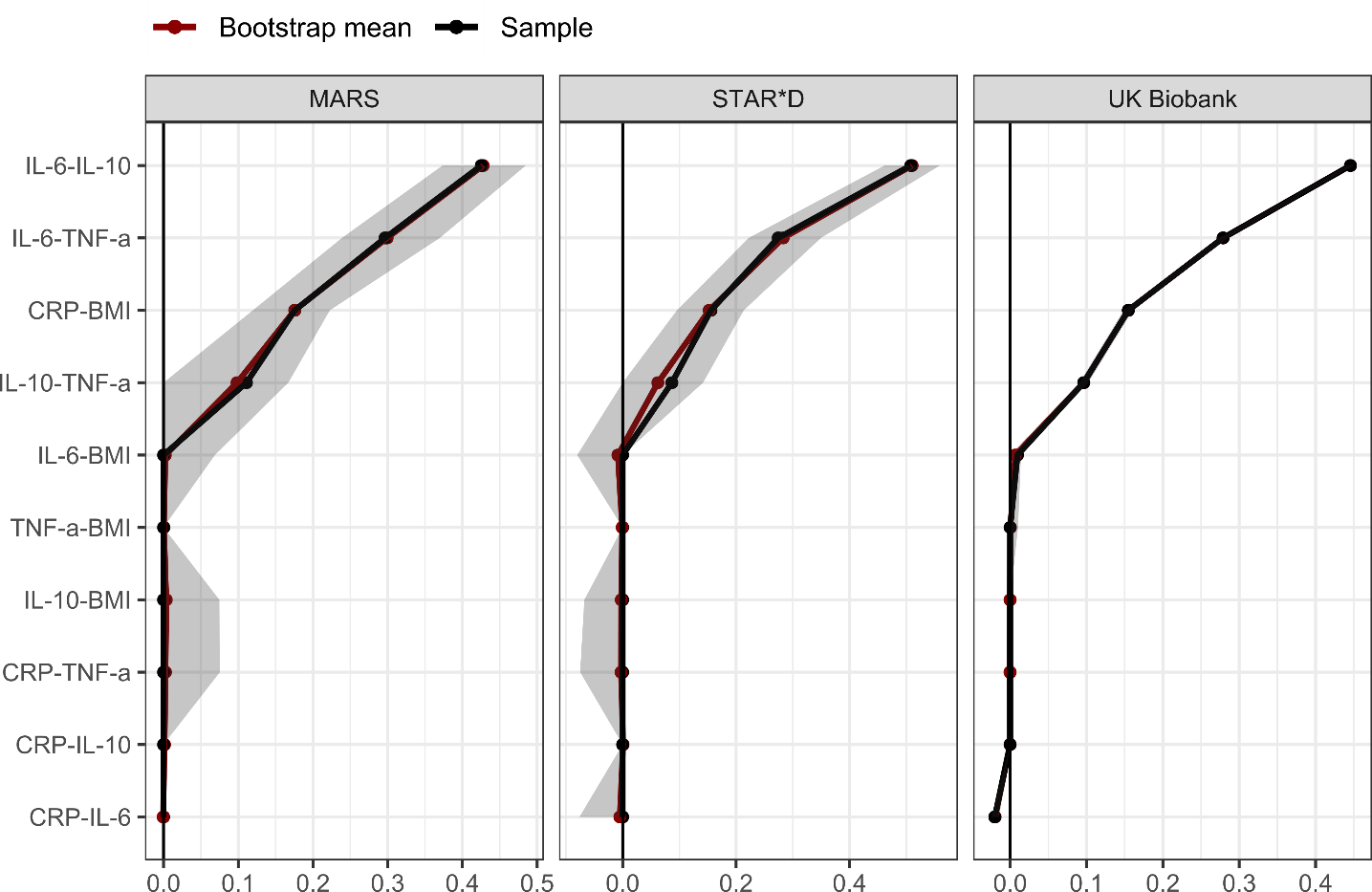

*Note*: Bootstrapped 95% quantile intervals (i.e., 95% of the distribution of raw bootstrapped edge estimates) are highlighted as shaded area for each edge. Black points indicate the raw FGL sample estimate while red points indicate the raw bootstrapped mean estimate. Edges are indicated on the y-axis and sorted by mean edge size across samples in descending order.
